# Systematic analysis of loss-of-function variants across MODY genes demonstrates gene- and NMD-specific effects and identifies NMD-escape *INS* variants as a novel cause of MODY

**DOI:** 10.1101/2025.09.07.25334427

**Authors:** Thomas W. Laver, Aparajita Sriram, Matthew N. Wakeling, Zeynep Şiklar, Oguzhan Kalyon, Andrew T. Hattersley, Michael N. Weedon, Sarah Flanagan, Elisa De Franco, Kevin Colclough, Kashyap A. Patel

## Abstract

**Aims/hypothesis:** Accurate interpretation of loss-of-function (LOF) variants in MODY genes is essential for diagnosis but remains challenging, particularly for variants predicted to escape nonsense-mediated decay (NMD). We aimed to systematically evaluate the pathogenicity of LOF variants, stratified by NMD-triggering and NMD-escape status, across all known MODY genes.

**Methods:** We analysed ultra-rare LOF variants (minor allele frequency <1 in 10,000) in 5171 individuals of European ancestry with suspected MODY, compared with 155,501 population-based controls from UK Biobank. LOF variants in *ABCC8, GCK, HNF1A, HNF4A, HNF1B, INS, KCNJ11, NEUROD1, PDX1*, and *RFX6* were classified as NMD-triggering or NMD-escape. We tested for gene-level enrichment in cases versus controls. For novel associations, we performed replication in additional MODY cases, assessed familial co-segregation, and undertook *in-silico* protein modelling.

**Results:** LOF variants were significantly enriched in all MODY genes except *ABCC8* and *KCNJ11*. Both NMD-triggering and NMD-escape variants were enriched in *GCK, HNF1A*, and *HNF4A*, consistent with haploinsufficiency (all *P* <10^-3^). *HNF1B* and *RFX6* showed enrichment only for NMD-triggering variants, while *NEUROD1* and *PDX1* were enriched for only NMD-escape variants.

A novel finding was significant enrichment of only NMD-escape LOF variants in *INS* (OR = 181, *P* < 10^-5^). Including replication, we identified eight families with 17 affected individuals carrying *INS* variants. These variants co-segregated with diabetes (LOD score = 3), included one *de novo* case, and were absent from >800,000 population controls. Individuals presented with diabetes at a median age of 19 years, had median BMI of 22.9 kg/m^2^, were negative for islet autoantibodies, and had low type 1 diabetes genetic risk scores. Compared with *INS* missense MODY, diagnosis occurred ∼10 years later. Protein modelling suggested that *INS* NMD-escape variants produce aberrant proinsulin molecules with unpaired B chain cysteines, leading to milder misfolding.

**Conclusions/interpretation:** The pathogenicity of LOF variants in MODY genes depends on gene context and NMD status. Heterozygous NMD-escape LOF variants in *INS* are a novel cause of MODY. These findings provide systematic gene-level evidence to inform variant interpretation guidelines and improve the accuracy of MODY diagnosis in clinical practice.

**Research in Context:** *What is already known about this subject?:*

- Interpretation of loss-of-function (LOF) variants is complex and strongly influenced by predicted NMD status.
- Current evidence for LOF variants in MODY genes is mainly based on case reports.
- Systematic genetic evidence is needed to improve diagnosis and to inform gene-specific variant interpretation guidelines.

*What is the key question?:*

- Do NMD-triggering and NMD-escape LOF variants contribute to MODY across all known MODY genes?

*What are the new findings?:*

- Heterozygous NMD-escape LOF variants in *INS* are a novel cause of MODY.
- Heterozygous LOF variants in *ABCC8* and *KCNJ11* are not enriched in MODY.
- Enrichment of NMD-triggering and NMD-escape LOF variants differs across MODY genes, clarifying underlying mechanisms and supporting gene-specific interpretation.

*How might this impact on clinical practice in the foreseeable future?:*

- These results improve the accuracy of MODY diagnosis by providing robust evidence for variant interpretation and supporting gene-specific clinical guidelines.

## Introduction

Accurate interpretation of genetic variants in MODY (maturity-onset diabetes of the young) genes is critical for diagnosis but remains a challenge in clinical practice. Variant classification remains challenging due to uncertainty in functional impact, the complexity of disease mechanisms, and evolving evidence. This is particularly true for loss-of-function (LOF) variants.

Interpreting LOF variants is complex because their impact on protein function and disease risk depends on their position within the gene. LOF variants typically include nonsense, frameshift, essential splice site changes, and exon deletions. LOF variants located more than 50 base pairs upstream of the final exon–intron junction typically trigger nonsense-mediated mRNA decay (NMD), resulting in haploinsufficiency. These are usually pathogenic when haploinsufficiency is a known disease mechanism. However, in the absence of gene-specific evidence, interpretation remains uncertain. In contrast, LOF variants in the non-NMD region, may avoid mRNA degradation and produce truncated proteins with unpredictable effects. These can retain function, result in complete loss of function, or act in a dominant-negative manner, making classification particularly challenging. (1) Although international guidelines exist for interpreting such variants, they are generic, and there is strong recommendation for the development of gene-specific guidance. (2,3)

Current MODY-specific guidelines for LOF variants covers *GCK, HNF1A*, and *HNF4A*, and further evidence is needed to classify these variants across all MODY genes. There is strong genetic evidence that NMD-triggering LOF variants in some MODY genes (for example, *GCK, HNF1A*, and *HNF4A*) cause disease via haploinsufficiency. However, for other MODY genes, such as *ABCC8, KCNJ11* and *INS*, systematic evidence is lacking. The diabetes phenotypes caused by *ABCC8* and *KCNJ11* are typically associated with activating missense variants, while for *INS* they are associated dominant negative missense variants (e.g., (4–6)), and NMD-triggering LOF variants would not be expected to be pathogenic. However, there are case reports of heterozygous LOF *ABCC8* variants in MODY, highlighting the need for further investigation (7).

Evidence for the pathogenicity of NMD-escape LOF variants in MODY is particularly limited. International guidelines recommend classifying NMD-escape LOF variants in *GCK, HNF1A* and *HNF4A* as pathogenic (8), but these recommendations are based on case reports rather than robust statistical genetic evidence (9–11). Isolated case reports also suggest that NMD-escape LOF variants in, *PDX1, HNF1B, RFX6* and *INS* may be disease-causing, but again, systematic evidence is lacking (12–15).

In this study, we perform a rare LOF variant burden analysis in known MODY genes, comparing individuals with clinically suspected MODY against large cohorts of population controls. By evaluating both NMD-triggering and NMD-escape LOF variants, we aim to clarify their pathogenicity, contribute to gene-specific interpretation guidelines, and ultimately improve the genetic diagnosis of MODY.

## Research Design and Methods

### Study Populations

#### MODY Cohort

We included 5,171 unrelated individuals of European ancestry with clinically suspected MODY, referred for genetic testing as part of routine clinical care through the Exeter Genomics Laboratory before October 2023. Clinical features of this cohort are summarised in Supplementary Table 1. The study was approved by the North Wales Research Ethics Committee (REC reference: 17/WA/0327), and all participants, or their legal guardians, provided written informed consent.

#### UK Biobank

We used the UK Biobank as a population-based control cohort. This prospective UK study recruited individuals aged 40–70 years irrespective of health status and collected extensive phenotypic and genetic data. For this analysis, we included participants from the 200,000 whole-genome sequencing release. All participants gave informed consent, and the UK Biobank Research Ethics Committee approved the use of these data (REC reference: 11/NW/0382).

#### gnomAD

For sensitivity analyses, we used version 3.1.2 of the Genome Aggregation Database (gnomAD), which comprises whole-genome sequencing data from 76,156 individuals (16). This version of GnomAD does not include UK Biobank participants.

#### Replication MODY Cohort for INS Loss-of-Function Variants and family member testing

To identify additional carriers of candidate *INS* variants, we reviewed two replication groups: (i) individuals of non-European ancestry with suspected MODY who underwent tNGS before October 2023 (n = 1,692), and (ii) all cases referred from the UK with suspected MODY between October 2023 and November 2024 (n = 505). To evaluate cosegregation, we tested family members for INS loss-of-function variants using Sanger sequencing.

### Genetic Analysis

#### MODY Cohort

We sequenced 2,571 individuals using targeted next-generation sequencing (tNGS) for all known MODY genes (17). The remaining 2600 individuals underwent Sanger sequencing for one or more of *GCK* (n = 941), *HNF1A* (n = 1,292), or *HNF4A* (n = 602). We have described the tNGS methods in detail in previous studies (17). Briefly, we used a custom Agilent SureSelect exon-capture panel (Agilent Technologies, Santa Clara, CA, USA) targeting known MODY genes. Sequencing was done on an Illumina NextSeq and NovoSeq sequencers (Illumina, San Diego, CA, USA). We called variants using the Genome Analysis Toolkit (GATK).

We carried out Sanger sequencing using BigDye Terminator v3.1 (Applied Biosystems). After removing unincorporated dye-terminators, we ran the fragments on an ABI 3730XL capillary DNA sequencer (Applied Biosystems). We called variants using Mutation Surveyor (SoftGenetics) and all included variants were manually checked using the electropherograms. We determined ancestry from tNGS data using an adapted version of the LASER method optimised for targeted panel data (18). For individuals without tNGS data, we used self-reported ethnicity.

We annotated variants using the following RefSeq transcripts: *ABCC8* (NM_000352.6), *GCK* (NM_000162.5), *HNF1A* (NM_000545.8), *HNF4A* (NM_175914.5), *HNF1B* (NM_000458.4), *INS* (NM_000207.3), *KCNJ11* (NM_000525.3), *NEUROD1* (NM_002500.4), *PDX1* (NM_000209.4), and *RFX6* (NM_173560.4). We performed annotation using Alamut Batch v1.11 (Interactive Biosoftware) on the GRCh37 genome build and converted coordinates to GRCh38 for comparison with control data.

We calculated logarithm of the odds (LOD) scores to assess co-segregation, following ClinGen guidelines (19). We used IBDseq (version r1206) to identify shared haplotypes between the *INS* LOF carriers (20).

#### UK Biobank

We analysed whole-genome sequencing data from 155,501 unrelated individuals of European ancestry. We chose genome sequencing over exome sequencing because it provides more uniform coverage across coding regions and reduces technical artefacts (21). Variant calling was performed on the GRCh38 reference genome, and we annotated variants using Alamut Batch v1.11. Szustakowski *et al*. (22) describe the sequencing methods in detail; further documentation is available at https://biobank.ctsu.ox.ac.uk/showcase/label.cgi?id=170.

#### gnomAD

We used data from 34,029 non-Finnish European individuals in gnomAD v3.1.2 as an additional control cohort. The gnomAD team used a BWA-Picard-GATK pipeline for joint variant calling and applied quality control using the Hail framework (16). They performed variant calling on GRCh38, and we annotated variants using Alamut Batch v1.11.

#### Definition of Loss-of-Function Variants

We defined loss-of-function (LOF) variants as stop-gain, essential splice site, or frameshift variants. Following published criteria (1), we classified LOF variants as NMD-escape if the premature termination codon lay in the final exon or within the last 50 base pairs of the penultimate exon. (Supplementary table 2) We classified all other LOF variants as NMD-triggering. For single exon genes like *KCNJ11* and *NEUROD1* variants are generally not expected to be susceptible to NMD, thus all LOF variants in these genes were analysed as NMD-escape variants.

#### Ultra-Rare Variant Burden Analysis

We tested for gene-level burden of ultra-rare LOF variants with a minor allele frequency (MAF) <1 in 10,000. We applied strict sample and variant-level quality control to minimise technical artefacts and included only high-quality variants across all cohorts (supplementary methods). We restricted the analysis to individuals of European ancestry to reduce population stratification.

We used Fisher’s exact test to compare the frequency of LOF variants between MODY cases and population controls, and we calculated odds ratios (ORs) with 95% confidence intervals (CIs). To check for test inflation and identify technical artefacts, we included synonymous variants as negative controls. We included *GCK, HNF1A*, and *HNF4A* as positive controls because they have ClinGen guidelines for interpreting NMD-triggering and NMD-escape variants (8).

To maximise power, we combined cases sequenced by Sanger and tNGS when analysing *GCK, HNF1A* and *HNF4A*. We first analysed all LOF variants together, then repeated analyses for NMD-triggering and NMD-escape variants separately. We corrected for multiple testing using a Bonferroni-adjusted significance threshold of *P* < 0.0025 (0.05 /10 genes × 2 variant categories: LOF and synonymous). Our analysis had 80% power to detect odds ratios of at least 8.9 for variants with a MAF of 1 in 10,000 in GCK, *HNF1A* and *HNF4A*, and an odds ratio of 9.8 for the other genes. (Supplementary table 13)

#### Sensitivity Analyses

We carried out several sensitivity analyses to test the robustness of our findings. We repeated burden tests using stricter (MAF <1 in 20,000) and more lenient (MAF <1 in 5,000) frequency thresholds to confirm that results were not influenced by the choice of frequency cutoff. We also repeated the analysis using gnomAD v3.1.2 as an alternative control cohort to confirm consistency.

#### Protein alignment and modelling

We used EMBL EBI’s Clustal Omega multiple sequence alignment program to generate alignments for predicted amino acid sequences in *INS* (23). We used AlphaFold Server (<u>AlphaFold Server -Google DeepMind</u>) to generate protein models and visualise them.

## Results

### Both NMD-triggering and NMD-escape LOF variants in HNF1A, HNF4A and GCK are enriched in the MODY cohort

The enrichment of rare variants in a disease cohort provides strong genetic evidence of pathogenicity (2). We therefore assessed the enrichment of ultra-rare (MAF <1 in 10,000) loss-of-function variants (stop-gain, essential splice site, or frameshift variants) in a MODY cohort compared to 155,501 controls. We first focused on the three most common MODY genes with robust evidence for causality for LOF variants (*GCK, HNF1A*, and *HNF4A*). We found no enrichment of ultra-rare synonymous variants in these genes, confirming that our analysis was well calibrated (Figure 1A, Supplementary table 3). In contrast, we observed strong enrichment of LOF variants in all three genes in the MODY cohort (all *P* < 10^−39^; Figure 1B).

**Figure 1:**
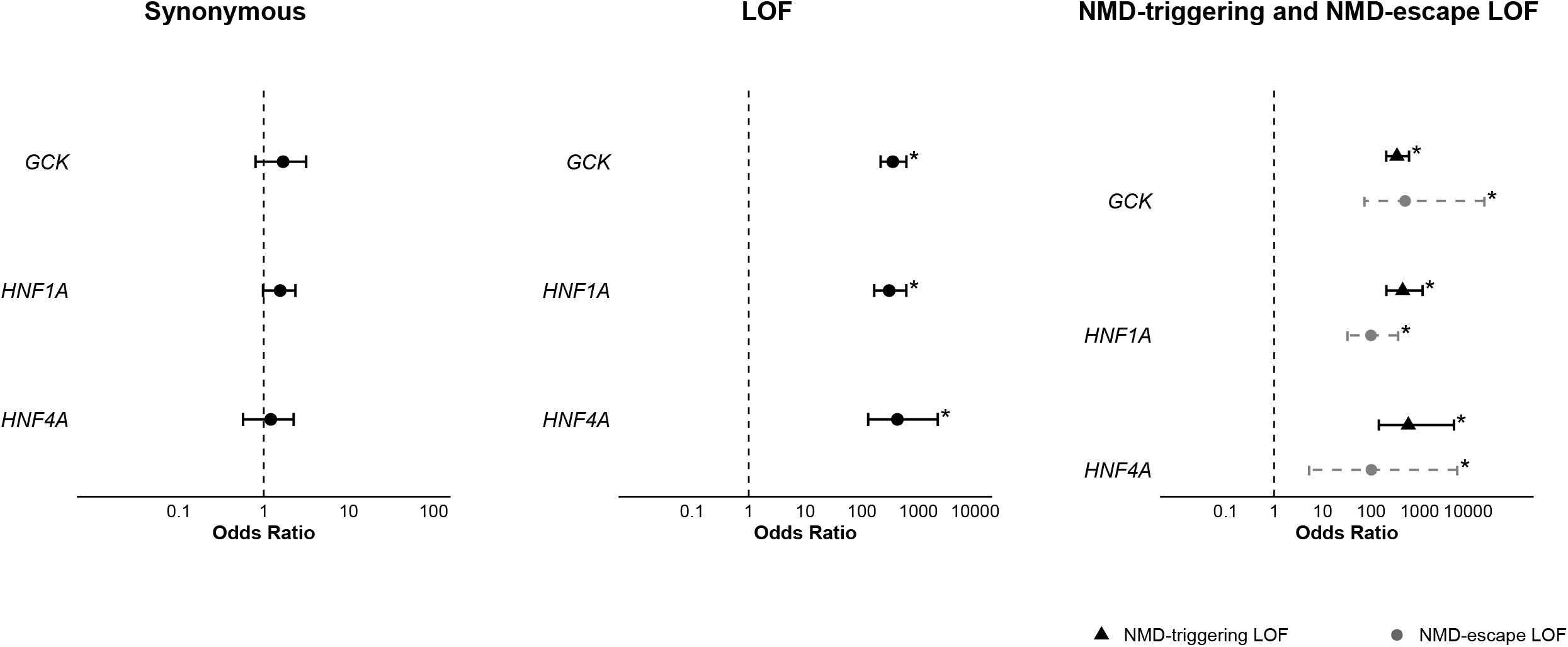
Gene-level burden analysis of ultra-rare variants in *GCK, HNF1A*, and *HNF4A* within the MODY cohort. This figure presents gene-level burden tests comparing ultra-rare variants (MAF < 1×10^−4^) between a European-ancestry MODY cohort (n = 2,471) and UK Biobank controls (n = 155,501). The analyses include: (A) synonymous variants; (B) protein-truncating variants (PTVs); and (C) PTVs further divided into those subject to nonsense-mediated decay (NMD) and those predicted to escape NMD. Asterisks (*) denote statistical significance after correction for multiple testing (p < 0.0025). Odds ratios and 95% confidence intervals are reported for each association. (Refer Supplementary Table 3)

To explore this association further, we analysed LOF variants predicted to trigger MND separately from those predicted to escape MND. As expected, the NMD-triggering variants, well-established causes of MODY, were strongly enriched in the MODY cohort (OR = 341, 441, 584 respectively for *GCK, HNF1A* and *HNF4A* respectively, all *P* < 10^−36^, Figure 1C), demonstrating that our analysis was powered to detect true associations.

We also observed marked enrichment of NMD-escape LOF variants in *GCK* (OR = 502, *P* < 10^−19^), *HNF1A* (OR = 99, *P* < 10^−19^), and *HNF4A* (OR = 101, *P* < 10^−3^) (Figure 1C, Supplementary table 3). The effect sizes for NMD-escape and NMD-triggering variants were similar for *GCK* and *HNF4A*, but slightly lower for *HNF1A* (interaction *P* = 0.046). Taken together, these findings provide strong support for the ClinGen recommendations to report LOF variants in *GCK, HNF1A*, and *HNF4A* as pathogenic when consistent with the clinical phenotype.

### LOF variants are not enriched in ABCC8 or KCNJ11, while enrichment in other MODY genes depends on NMD status

We next applied our analysis to the remaining MODY genes, where the role of LOF variants is less well established and the relevance of NMD-escape versus NMD-triggering variants is unclear. We found no enrichment of ultra-rare synonymous variants in any of these genes, confirming that our analysis was well calibrated (Figure 2A, Supplementary table 4).

**Figure 2:**
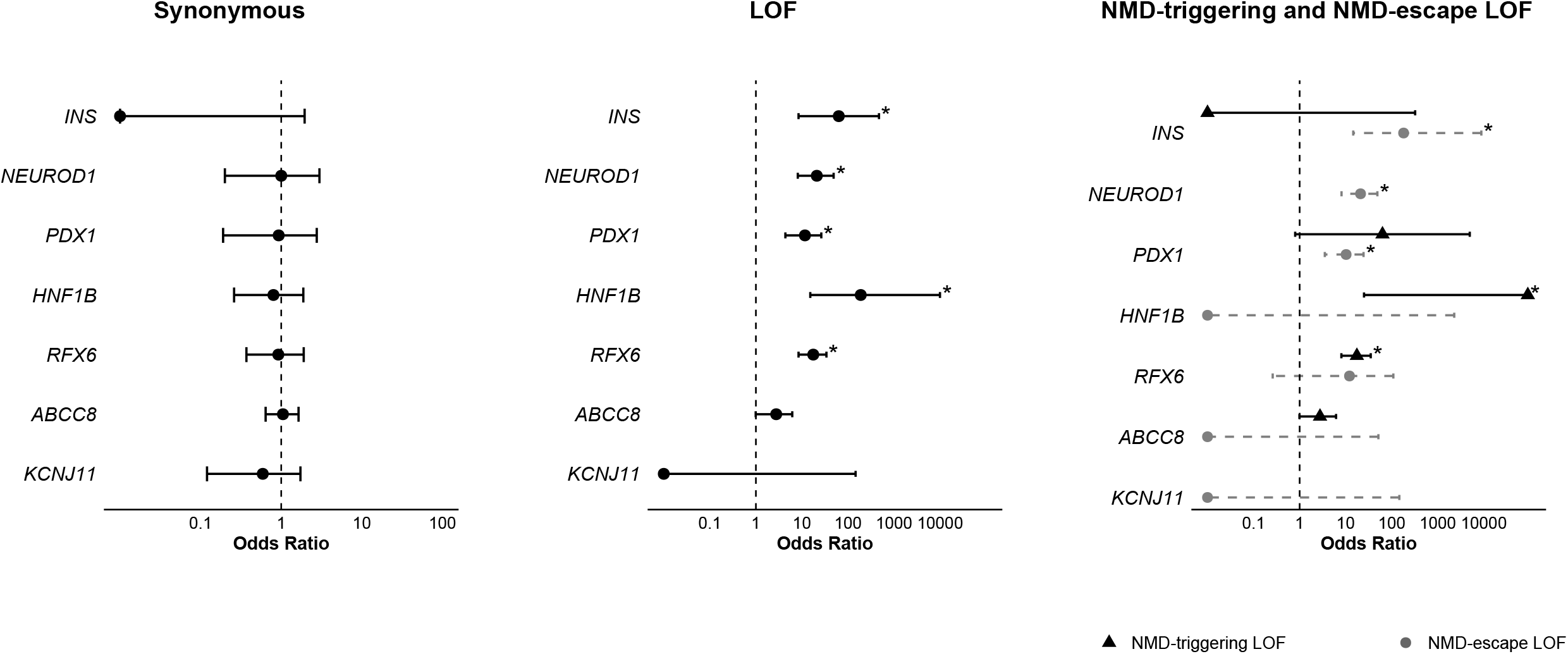
Gene-level burden analysis of ultra-rare variants in MODY genes within the MODY cohort. This figure presents gene-level burden tests comparing ultra-rare variants (MAF < 1×10^−4^) between a European-ancestry MODY cohort (n = 2,471) and UK Biobank controls (n = 155,501) in MODY genes *INS, NEUROD1, PDX1, HNF1B, RFX6, ABCC8* and *KCNJ11*. The analyses include: (A) synonymous variants; (B) protein-truncating variants (PTVs); and (C) PTVs further divided into those subject to nonsense-mediated decay (NMD) and those predicted to escape NMD. Asterisks (*) denote statistical significance after correction for multiple testing (p < 0.0025). Odds ratios and 95% confidence intervals are reported for each association. (Refer Supplementary Table 4)

We first looked at all LOF variants irrespective of the NMD status. We found lack of enrichment in *KCNJ11* (OR = 0, *P* > 1) or *ABCC8* (OR = 2.8, *P* > 0.03) (Figure 2B, Supplementary table 4). In contrast, we observed significant enrichment of LOF variants in *HNF1B* (OR = 189, *P* < 10^−4^), *RFX6* (OR = 18, *P* < 10^−10^), *PDX1* (OR = 12, *P* < 10^−5^), *NEUROD1* (OR = 21, *P* < 10^−7^), and, notably, *INS* (OR = 63, *P* < 10^−4^).

We then split LOF variants by predicted NMD status to assess whether the observed associations differed. As *NEUROD1* and *KCNJ11* are single-exon genes, all LOF variants are expected to escape NMD, so separate analyses were not required. In the remaining genes, *ABCC8* showed no enrichment for either class (NMD-triggering: OR = 2.8, P > 0.03; NMD-escape: OR = 0, P = 1). By contrast, several other MODY genes displayed distinct patterns, with enrichment driven specifically by NMD-triggering LOF variants in *HNF1B* (OR = ∞, *P* < 10^−5^) and *RFX6* (OR = 17, *P* < 10^−11^), whereas NMD-escape variants were not enriched in either gene (P > 0.09). Conversely, the enrichment in *PDX1* (OR = 10, *P* < 10^−4^) and *INS* (OR = 180, *P* < 10^−5^) was driven primarily by NMD-escape variants. The *PDX1* results are consistent with previous literature whereas this is particularly notable for *INS*, as it provides the first large-scale evidence supporting NMD-escape *INS* variants as a novel cause of MODY.

Sensitivity analyses using different frequency thresholds (MAF <1 in 20,000, <1 in 5,000, or any frequency) and an alternative control cohort (gnomAD v3.1.2) produced consistent results (Supplementary Tables 6-9), supporting the robustness of these findings. Taken together, these results do not support a causal role for LOF variants in *ABCC8* or *KCNJ11*. However, they highlight a novel and potential pathogenic role for NMD-escape *INS* variants in MODY.

### Heterozygous NMD-escape LOF variants in INS identified in additional MODY cases, show strong familial co-segregation and are absent from population databases

Following the novel observation of enrichment of NMD-escape LOF variants in *INS*, we carried out further analyses to assess the strength of genetic evidence for pathogenicity at the variant level.

In our primary cohort, we identified three probands carrying p.Gln78* NMD-escape LOF variant in *INS*. To find additional cases, we reviewed all individuals of non-European ancestry with suspected MODY (n = 1,692) and recent referrals to our laboratory for genetic testing. This search identified five additional probands: two with the same p.Gln78* variant and three with other truncating variants: p.Leu82GlyfsTer52, p.Glu83ValfsTer58, and p.Cys95*. (Figure 3) In total, we identified eight families with NMD-escape *INS* variants, comprising 17 affected individuals (Figure 3).

**Figure 3:** Pedigrees of patients with NMD-escape loss-of-function (LoF) variants in the INS gene within the MODY cohort. Seven families carrying three different variants are shown, with Families 1-5 sharing the same p.Gln78* variant. Squares represent males, circles represent females. Black shading indicates individuals diagnosed with diabetes. Arrows mark the probands. Labels indicate variant status: N/M denotes tested and heterozygous for the variant; N/N denotes tested and does not carry the variant. Additional annotations include: Dx (age at diagnosis), Rx (treatment), Dur (duration of diabetes).

None of these variants were present in gnomAD v4 (n = 807,162), supporting their rarity in the general population. We confirmed that the p.Leu82GlyfsTer52 variant occurred *de novo* in the proband (Figure 3, Family 6). We had sufficient genotype data to assess familial co-segregation for the p.Gln78* variant. This variant showed strong co-segregation with diabetes, with a combined LOD score of 3.0 across five families (Figure 3, Families 1–5), consistent with full penetrance in the individuals tested. We assessed five families with p.Gln78* who had suitable available sequencing data to assess a shared haplotype, but none was detected. Taken together, these findings provide both variant-level and gene-level evidence supporting NMD-escape loss-of-function variants in *INS* as a novel cause of MODY.

### Cases with INS NMD-escape LOF variants have a phenotype consistent with a monogenic aetiology

We assessed whether the clinical features of individuals with *INS* NMD-escape PTVs supported a monogenic cause of diabetes. These individuals had a median age at diagnosis of 19 years, a median HbA1c of 7.9%, and a median BMI of 22.9 kg/m^2^ (Table 1). They also lacked islet autoantibodies associated with type 1 diabetes, and their genetic risk for type 1 diabetes was low (median first percentile of the type 1 diabetes population). They also showed evidence of residual insulin secretion, with a median C-peptide of 430 at a median diabetes duration of 13 years. In line with this, only 70.6% were treated with insulin. Together, these features are not consistent with a polygenic aetiology and support the causal role of *INS* NMD-escape LOF variants for MODY. Interesting, their clinical features were similar to the cases with pathogenic *INS* missense variants causing MODY (variants listed in Supplementary table 11) except *INS* NMD-escape LOF variant carriers were diagnosed 10 years later and had directionally higher random c-peptide. (Table 1) These data suggest that these variants are potentially less severe than pathogenic *INS* missense variants.

**Table 1:** Clinical features of MODY patients with NMD-escape LoF variants in *INS* compared to MODY patients with missense variants in *INS*. Median (IQR) for continuous variable and % for categorical variable. List of missense variants
provided in Supplementary Table 11.

|  | *INS* NMD-escape LOF<br>variant carriers (n=17) | *INS* missense variant<br>carriers (n= 43) | *P* value |
| --- | --- | --- | --- |
| Age of diagnosis (y) | 19(17-27) | 9(2-14) | 9.3 x 10<sup>-6</sup> |
| Duration diabetes (y) | 12.9(1.3-20.3) | 11.9(1.9-29.3) | 0.5 |
| Females | 70.6% | 46.5% | 0.0004 |
| BMI (kg/m<sup>2</sup>) | 22.9(20.4-27) | 22.2(19.8-26.2) | 0.6 |
| Parent diabetes | 93.8% | 71.9, 32% | 0.1 |
| HbA1C (%) | 7.9(6.5-10.5) | 7.9(7-12) | 0.9 |
| C-peptide (pmol/l) | 429.5 (150.0-1119.25), 14 | 300 (115.3-554.8) | 0.4 |
| Islet autoantibody positive | 0% | 5% | 1 |
| T1DGRS (centile) | 1.2 (0.7-16.8) | 5 (1.5-15.7) | 0.05 |
| Current insulin treatment | 70.6% | 90.2% | 0.1 |

### MODY-associated LOF INS variants potentially lead to milder misfolding

To investigate the mechanisms by which NMD-escape loss-of-function variants in *INS* cause MODY, we performed sequence alignment and structural modelling of our newly identified variants alongside all published LOF variants.

Correct folding of insulin requires precise disulphide bonding between cysteines in the B and A chains. All stop-gain MODY variants truncate the entire A chain, leaving the B chain cysteines unpaired and incapable of forming disulphide bonds (Figure 4, Supplementary figure 3). Similarly, MODY frameshift variants uniformly alter the reading frame by +2, abolishing the native A chain and extending translation into the 3⍰ UTR where all terminate. Although these frameshifts introduce one or two cysteines, structural modelling indicates that the new residues are potentially positioned too far from the B chain cysteines to form aberrant disulphide bonds (range 6.9-20.4 Å; typical distance for wild type *INS* ∼ 4.8 Å; Supplementary Figure 3). (24,25)

**Figure 4:**
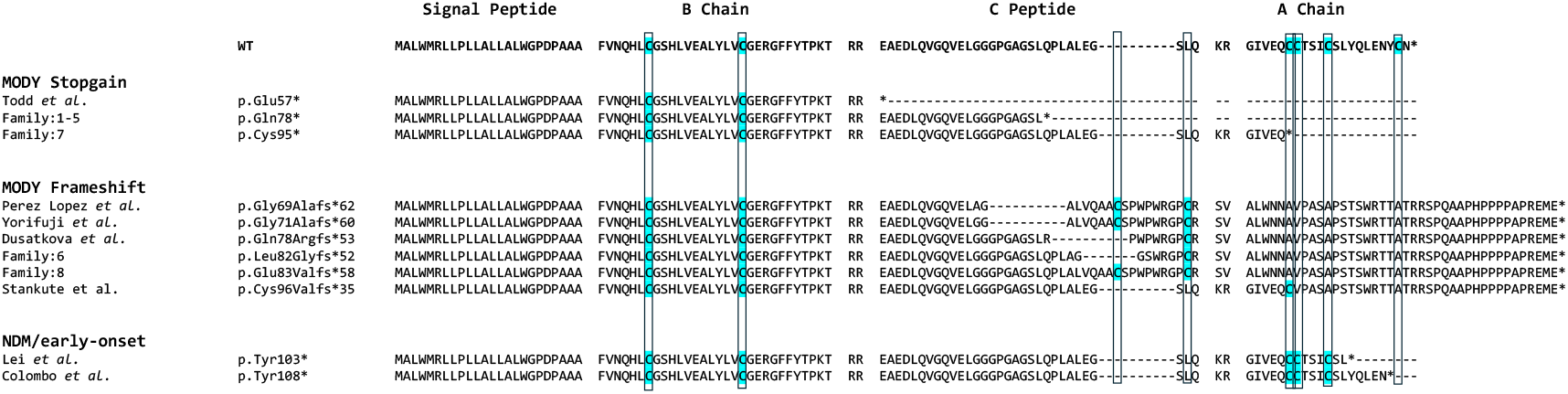
Predicted protein sequences of *INS* NMD-escape loss-of-function (LOF) variants. The wild-type *INS* sequence is shown at the top for reference. MODY-associated stop-gain and frameshift NMD-escape LOF variants identified in our cohort or reported in the literature are included. Published NMD-escape LOF variants linked to neonatal diabetes (NDM) or early-onset diabetes are also shown. Conserved cysteine residues are highlighted in blue.

These data indicate that LOF MODY variants consistently result in unpaired B chain cysteines. This appears to cause relatively modest misfolding, sufficient to activate the unfolded protein response but not at the intensity observed in neonatal diabetes. Consistent with this interpretation, no heterozygous frameshift variants have been reported in neonatal diabetes. By contrast, only two heterozygous truncating variants have been described, and both were NMD-escape variants (p.Tyr103*, p.Tyr108* (26,27)). These variants truncate late in the A chain, preserving the first three cysteines but removing the fourth. This configuration is predicted to permit aberrant crosslinking of B-chain cysteines, leading to severe misfolding and a stronger unfolded protein response (Supplementary Figure 3).

## Discussion

In this study, we provide gene-level evidence for the pathogenicity of both NMD-triggering and NMD-escape loss-of-function (LOF) variants across all known MODY genes. We show that the pathogenic impact of a LOF variant depends on whether it triggers NMD, and that this varies by gene. Specifically, we demonstrate that NMD-escape LOF variants in *INS* are a novel cause of MODY.

Our study provides the first systematic evidence that heterozygous NMD-escape LOF variants in *INS* cause MODY. This conclusion is supported by strong gene- and variant-level data. We observed significant enrichment in MODY cases compared with population controls (odds ratio = 181, *P* < 10^-5^), clear co-segregation in families (LOD score = 3.0), and one *de novo* occurrence. In total, we identified four variants in seven families, none of which are present in population databases comprising 807,162 individuals. Clinical features of affected individuals were consistent with monogenic diabetes and resembled those seen in patients with pathogenic MODY causing *INS* missense variants, though diagnosis occurred on average ten years later. Six previously published NMD-escape *INS* variants have been described in diabetes cases (Supplementary Table 11, Supplementary Figure 1). Three were reported as part of larger cohorts without detailed phenotyping (28–30), while three were described in individual case reports (13,31,32). Reported patients were diagnosed between 7 and 36 years of age (mean 20 years), consistent with our cohort. Together, this evidence establishes heterozygous *INS* NMD-escape LOF variants as a cause of MODY that should be reported in diagnostic testing. However, the lack of enrichment of NMD-triggering variants in our study and lack of diabetes in parents of recessive neonatal diabetes due to NMD-triggering variants suggest that variants that cause haploinsufficiency do not cause MODY and should not be reported. (33)

We hypothesise that NMD-escape LOF variants in *INS* act by generating aberrant proinsulin molecules with unpaired B chain cysteines. Variants with an intact A chain but loss of at least one cysteine allow abnormal disulphide bond formation, producing severe misfolding and neonatal diabetes. In contrast, variants that remove or disrupt the entire A chain leave unpaired B chain cysteines, leading to milder misfolding. (Supplementary figure 3) Our findings are consistent with previous *in vitro* work showing that truncated *INS* containing only the B chain fails to interact with wild-type insulin. Together, these data support a model in which MODY-associated variants are dominant negative, cause milder misfolding without toxic gain-of-function, resulting in a weaker unfolded protein response and slower β-cell loss. This mechanism remains a hypothesis that requires confirmation in cellular and *in vivo* systems.

Our results also clarify the role of LOF variants in *ABCC8* and *KCNJ11*. We found no evidence of enrichment, questioning the role of heterozygous LOF variants in these genes in MODY. This will need further follow up variant level genetic evidence to conclusively refute the causal role of these variants in MODY. Our findings however are consistent with known disease mechanisms of these genes in which activating variants in *ABCC8* and *KCNJ11* cause neonatal diabetes and MODY, while recessive LOF variants cause congenital hyperinsulinism (5,34). Thus, haploinsufficiency is not the underlying mechanism for MODY.

We identified additional gene-specific patterns. NMD-escape loss-of-function variants in *PDX1* were associated with MODY. We observed three frameshift variants disrupting the homeodomain, which is encoded by the second and last exon, and three frameshift/stop-gain variants occurring after the homeodomain; none of these cases carried other known pathogenic variants. These findings are consistent with a dominant-negative mechanism previously described (35). We were underpowered to assess NMD-triggering protein-truncating variants in isolation, as only one such case was present in the MODY cohort (p.Cys18*, unsolved female, diagnosed in their teens), providing an odds ratio of 63 (0.8-4941), *P* = 0.03. These results suggest that further variant-level evidence and larger studies are needed to clarify the role of NMD-triggering variants in *PDX1*. For *HNF1B* and *RFX6*, enrichment was confined to NMD-triggering LOF variants, with no evidence supporting pathogenicity of NMD-escape variants. Although isolated *RFX6* NMD-escape variants have been reported (36), our results suggest these should be interpreted cautiously. In contrast, we found enrichment of both NMD-triggering and NMD-escape protein-truncating variants in *GCK, HNF1A*, and *HNF4A*, consistent with ClinGen recommendations for reporting such variants. The presence of functional domains in the NMD-escape regions of *GCK* and *HNF1A* supports this mechanism, although *HNF4A* lacks a defined functional domain in the NMD-escape region, and the basis of pathogenicity remains to be determined. Overall, our results provide mechanistic insights into MODY gene action.

Our study has limitations. Although we leveraged one of the largest available MODY cohorts, we had power only to detect associations with odds ratios ≥ 9.8 at an allele frequency of 0.0001. We cannot exclude the possibility that other genes harbour ultra-rare or low-penetrance LOF variants undetected in our analysis. Our burden testing was limited to individuals of European ancestry, where statistical power was greatest. Future work should assess these associations across diverse ancestries. We used publicly available population controls, introducing differences in sequencing platforms; however, synonymous variant analyses support the robustness of our variant quality control measures.

In conclusion, we provide strong genetic evidence that heterozygous NMD-escape LOF variants in *INS* are a novel cause of MODY. Our systematic analysis across all MODY genes provides robust statistical evidence that can guide gene-specific variant interpretation, support ClinGen recommendations, and improve diagnosis of monogenic diabetes.

## Supporting information

Supplemental methods and results

## Data Availability

UK Biobank data is available at https://www.ukbiobank.ac.uk/enable-your-research and is accessible through application. GnomAD data is freely available to all at https://gnomad.broadinstitute.org/. The MODY cohort data is not publicly available for ethical and patient confidentiality related reasons but is available upon reasonable request to the corresponding author.

## Abbreviations

LOF: Loss of function
MODY: Maturity onset diabetes of the young
NMD: Nonsense-mediated decay.

## Acknowledgments

This research has been conducted using the UK Biobank Resource. This work was conducted under UK Biobank project number 9072. KAP is funded by the Wellcome Trust (219606/Z/19/Z). T.W.L is supported by the Academy of Medical Sciences/the Wellcome Trust/the Government Department of Science Innovation and Technology/the British Heart Foundation/Diabetes UK Springboard Award [SBF009\1135]. ATH is supported by Wellcome Trust Senior Investigator award (WT098395/Z/12/Z). The work is supported by the National Institute for Health Research (NIHR) Exeter Biomedical Research Centre, Exeter, UK. And NIHR Exeter Clinical Research Facilities. The Wellcome Trust, MRC and NIHR had no role in the design and conduct of the study; collection, management, analysis, and interpretation of the data; preparation, review, or approval of the manuscript; and decision to submit the manuscript for publication. The views expressed are those of the author(s) and not necessarily those of the Wellcome Trust, Department of Health, NHS or NIHR. For the purpose of open access, the author has applied a CC BY public copyright licence to any Author Accepted Manuscript version arising from this submission.

## Authors’ relationships and activities

The authors declare no competing interests.

## Contribution Statement

T.W.L. and A.S. are listed as joint first authors. The study was conceptualised by K.A.P., M.N.We., and T.W.L., with methodology contributions from K.A.P., M.N.Wa., M.N.We., and T.W.L. A.S. performed the investigation, formal analysis, and visualization. Resources were provided by A.T.H., E.D.F., K.C., O.K., S.F., and Z.S. The original draft was prepared by A.S., K.A.P., and T.W.L., with A.S., A.T.H., E.D.F., K.C., K.A.P., M.N.Wa., M.N.We., O.K., S.F., T.W.L., and Z.S. contributing to review and editing. Supervision was provided by A.T.H. and K.A.P., and funding acquisition was undertaken by K.A.P., M.N.We., and T.W.L. K.A.P. is the guarantor of this work and, as such, had full access to all the data in the study and takes responsibility for the integrity of the data and the accuracy of the data analysis.

