## Supplemental methods and results for "Systematic analysis of loss-of-function variants across MODY genes demonstrates gene- and NMD-specific effects and identifies NMD-escape *INS* variants as a novel cause of MODY"

**Supplementary Methods**

**Quality control**

**MODY cohort**

For our tNGS sequenced samples, we performed a rigorous quality control (QC) process at the sample, variant, and genotype levels to ensure the accuracy and consistency of sequencing data obtained from different platforms. Recognizing the potential for discrepancies between sequencing technologies, we applied stringent filters to minimize technical artifacts and enhance reliability. We began by excluding six samples that had a genotype missingness rate greater than 2%.

We then applied multiple filters at the variant level. We removed variants with a strand bias FS score greater than 60, a quality-by-depth (QD) score less than 2, a ReadPosRankSum score below -8, or a MQRankSum score below -12.5. We also excluded variants with a mapping quality less than 40, a read depth below 20, or a genotype quality below 20. To account for allelic imbalance, we removed variants for which a binomial test yielded a p-value less than 0.001. We additionally excluded variants located in the *HNF1A* c-insertion region (chr12:120994310-120994335), which is known to produce false positives. Finally, we removed any genotypes with a missingness rate greater than 2%.

After applying all quality control steps, we retained 554 high-quality variants for downstream analyses.

**UK Biobank**

We applied the same quality control strategy to the UK Biobank dataset, with minor adjustments to reflect its use of whole-genome sequencing rather than targeted gene panels. Specifically, we lowered the read depth threshold to 15 for variant exclusion. We also removed variants with an AAscore below 0.5, a metric generated by Graphtyper that estimates the likelihood of a variant being a true positive. (1) Furthermore, we excluded any variants located in low-complexity regions as flagged by gnomAD. To maintain consistency between cohorts, we excluded any variant that failed quality control in either the MODY cohort or the UK Biobank from both datasets.

**gnomAD**

We used publicly available gnomAD data, which had already undergone sample and variant-level quality control. In addition to these existing filters, we excluded variants that were located in regions with low coverage, defined as ≤10× in more than 20% of samples. We also removed variants that were filtered by gnomAD or flagged as being located in low-complexity regions. As with the MODY and UK Biobank datasets, we removed any variant that failed quality control in either the MODY cohort or gnomAD from both datasets prior to analysis.

**Supplementary Results**

**Supplementary table 1: Characteristics of MODY cohort**

| Characteristics | MODY cohort |
| --- | --- |
| N | 5,171 |
| Age of diagnosis of diabetes, years | 22 (15-20) |
| Female Sex, % | 59.2 |
| Age at recruitment, years | 29.6 (18.9-38.6) |
| BMI, kg/m^2^ | 25.2 (22.1-29.6) |
| Parents with diabetes, % | 69.4 |
| HbA1c, % | 7.3 (6.4-9.3) |
| On Insulin treatment, % | 45.7 |

Median (IQR) for continuous variable and n (%) for categorical data

**Supplementary table 2: NMD-escape regions in each MODY gene from Ensembl (GRCh38)**

| Gene | Transcript | No. of exons | Direction | Chr | Last exon | | Last 50 bp of penultimate exon | |
| --- | --- | --- | --- | --- | --- | --- | --- | --- |
|  |  |  |  |  | start | end | start | end |
| *INS* | NM_000207.3 | 2 (+1 noncoding) | 3'>5' | 11 | 2159997 | 2159852 | 2160835 | 2160785 |
| *PDX1* | NM_000209.4 | 2 | 5'>3' | 13 | 27924256 | 27924701 | 27920494 | 27920544 |
| *HNF1A* | NM_000545.8 | 10 | 5'>3' | 12 | 121001065 | 121001192 | 120999577 | 120999627 |
| *HNF4A* | NM_175914.5 | 10 | 5'>3' | 20 | 44429524 | 44429665 | 44428437 | 44428487 |
| *GCK* | NM_000162.5 | 10 | 3'>5' | 7 | 44145280 | 44145136 | 44145547 | 44145497 |
| *RFX6* | NM_173560.4 | 19 | 5'>3' | 6 | 116931331 | 116931506 | 116928921 | 116928971 |
| *ABCC8* | NM_000352.6 | 39 | 3'>5' | 11 | 17393128 | 17392991 | 17393747 | 17393697 |
| *HNF1B* | NM_000458.4 | 9 | 3’>5’ | 17 | 37687392 | 37687372 | 37699126 | 37699076 |

**Supplementary table 3: Gene burden tests for *GCK*, *HNF1A*, *HNF4A* in the MODY cohort (n=5,171) with UK biobank (n=155,501) as controls**

| Variant type | Gene | Allele count in MODY cohort | Allele count in population cohort  (UK biobank) | P value | Odds ratio | 95% CI lower limit | 95% CI upper limit |
| --- | --- | --- | --- | --- | --- | --- | --- |
| NMD-triggering PTV  (MAF < 0.0001) | *GCK* | 125 | 17 | 2.78 x10^-188^ | 341.34 | 204.69 | 604.84 |
|  | *HNF1A* | 74 | 7 | 8.92 x10^-112^ | 441.18 | 203.69 | 1135.62 |
|  | *HNF4A* | 23 | 2 | 1.13 x10^-37^ | 584.1 | 144.26 | 5110.68 |
| NMD-triggering Synonymous (MAF < 0.0001) | *GCK* | 8 | 236 | 0.3 | 1.55 | 0.66 | 3.1 |
|  | *HNF1A* | 18 | 517 | 0.2 | 1.44 | 0.85 | 2.3 |
|  | *HNF4A* | 9 | 358 | 0.4 | 1.27 | 0.58 | 2.44 |
| NMD-escape PTV  (MAF < 0.0001) | *GCK* | 11 | 1 | 5.22x x10^-18^ | 502.13 | 72.95 | 21609.47 |
|  | *HNF1A* | 12 | 5 | 5.43x10^-18^ | 99.33 | 32.56 | 359.97 |
|  | *HNF4A* | 2 | 1 | 0.001 | 101.24 | 5.27 | 5972.06 |
| Non-NMD region: Synonymous (MAF < 0.0001) | *GCK* | 2 | 34 | 0.2 | 2.68 | 0.31 | 10.46 |
|  | *HNF1A* | 5 | 92 | 0.08 | 2.25 | 0.71 | 5.44 |
|  | *HNF4A* | 1 | 61 | 1 | 0.83 | 0.02 | 4.79 |

2,571 individuals had tNGS. 2,600 individuals underwent Sanger sequencing for either ***GCK*** (n = 941), ***HNF1A*** (n = 1,292), or ***HNF4A*** (n = 602).

**Supplementary table 4: Gene burden test (MAF < 0.0001) in MODY cohort (n=2,571) with UK biobank (n=155,501) as controls**

| Variant type | Gene | Allele count in MODY cohort | Allele count in population cohort  (UK biobank) | P value | Odds ratio | 95% CI lower limit | 95% CI upper limit |
| --- | --- | --- | --- | --- | --- | --- | --- |
| NMD-triggering PTV | *ABCC8* | 6 | 131 | 0.03 | 2.77 | 0.99 | 6.19 |
|  | *HNF1B* | 3 | 0 | 4.31x10^-6^ | Inf | 24.98 | Inf |
|  | *INS* | 0 | 2 | 1 | 0.00 | 0.00 | 322.22 |
|  | *PDX1* | 1 | 1 | 0.03 | 62.94 | 0.8 | 4940.62 |
|  | *RFX6* | 11 | 38 | 3.55 x10^-10^ | 17.49 | 8.06 | 34.94 |
| NMD-escape PTV | *ABCC8* | 0 | 6 | 1 | 0 | 0 | 51.28 |
|  | *HNF1B* | 0 | 1 | 1 | 0 | 0 | 2273.75 |
|  | *INS* | 3 | 1 | 1.70x10 x10^-5^ | 180.88 | 14.57 | 8809.85 |
|  | *KCNJ11* | 0 | 3 | 1 | 0 | 0 | 146.11 |
|  | *NEUROD1* | 8 | 23 | 2.80x10^-8^ | 21.02 | 8.13 | 48.72 |
|  | *PDX1* | 6 | 37 | 0.0001 | 10.22 | 3.52 | 24.42 |
|  | *RFX6* | 1 | 5 | 0.09 | 12.09 | 0.26 | 108.27 |
| NMD-triggering synonymous | *ABCC8* | 20 | 1110 | 0.64 | 1.09 | 0.66 | 1.69 |
|  | *HNF1B* | 5 | 378 | 0.84 | 0.8 | 0.26 | 1.88 |
|  | *INS* | 0 | 47 | 1 | 0 | 0 | 4.94 |
|  | *PDX1* | 2 | 93 | 0.67 | 1.3 | 0.16 | 4.83 |
|  | *RFX6* | 6 | 440 | 0.85 | 0.82 | 0.3 | 1.81 |
| NMD-escape synonymous | *ABCC8* | 0 | 36 | 1 | 0 | 0 | 6.52 |
|  | *HNF1B* | 0 | 0 | 1 | 0 | 0 | Inf |
|  | *INS* | 0 | 69 | 0.63 | 0 | 0 | 3.32 |
|  | *KCNJ11* | 3 | 309 | 0.5 | 0.59 | 0.12 | 1.73 |
|  | *NEUROD1* | 3 | 181 | 1 | 1.00 | 0.20 | 2.98 |
|  | *PDX1* | 1 | 102 | 1 | 0.59 | 0.01 | 3.38 |
|  | *RFX6* | 1 | 22 | 0.31 | 2.75 | 0.07 | 16.99 |

**Supplementary table 5: Sensitivity analysis gene burden test in MODY Sanger Sequencing cohort (n=2,600) with UK biobank (n=155,501) as controls**

| Variant type | Gene | Allele count in MODY cohort | Allele count in population cohort  (UK biobank) | P value | Odds ratio | 95% CI lower limit | 95% CI upper limit |
| --- | --- | --- | --- | --- | --- | --- | --- |
| NMD-triggering PTV (All) | *GCK* | 102 | 17 | 6.56 x10^-206^ | 991.82 | 588.36 | 1771.38 |
|  | *HNF1A* | 39 | 7 | 3.54 x10^-74^ | 670.78 | 296.23 | 1777.76 |
|  | *HNF4A* | 9 | 2 | 1.08 x10^-20^ | 1161.48 | 239.90 | 11048.85 |
| NMD-triggering PTV  (MAF < 0.0001) | *GCK* | 102 | 17 | 6.56 x10^-206^ | 991.82 | 588.36 | 1771.38 |
|  | *HNF1A* | 39 | 7 | 3.54 x10^-74^ | 670.78 | 296.23 | 1777.76 |
|  | *HNF4A* | 9 | 2 | 1.08 x10^-20^ | 1161.48 | 239.90 | 11048.85 |
| NMD-escape PTV (All) | *GCK* | 10 | 1 | 6.95 x10^-22^ | 1653.03 | 234.84 | 71724.87 |
|  | *HNF1A* | 7 | 5 | 1.99 x10^-12^ | 168.55 | 46.01 | 673.80 |
|  | *HNF4A* | 2 | 1 | 4.46 x10^-5^ | 516.63 | 26.86 | 30475.23 |
| NMD-escape PTV  (MAF < 0.0001) | *GCK* | 10 | 1 | 6.95 x10^-22^ | 1653.03 | 234.84 | 71724.87 |
|  | *HNF1A* | 7 | 5 | 1.99 x10^-12^ | 168.55 | 46.01 | 673.80 |
|  | *HNF4A* | 2 | 1 | 4.46 x10^-5^ | 516.63 | 26.86 | 30475.23 |

2,600 individuals underwent Sanger sequencing for either ***GCK*** (n = 941), ***HNF1A*** (n = 1,292), or ***HNF4A*** (n = 602).

| Variant type | Gene | Allele count in MODY cohort | Allele count in population cohort  (UK biobank) | P value | Odds ratio | 95% CI lower limit | 95% CI upper limit |
| --- | --- | --- | --- | --- | --- | --- | --- |
| NMD-triggering PTV | *ABCC8* | 4 | 102 | 0.1 | 2.37 | 0.63 | 6.26 |
|  | *HNF1B* | 3 | 0 | 4.31x10^-6^ | Inf | 24.98 | Inf |
|  | *INS* | 0 | 2 | 1 | 0 | 0 | 322.22 |
|  | *PDX1* | 1 | 1 | 0.03 | 62.94 | 0.8 | 4940.62 |
|  | *RFX6* | 11 | 38 | 3.55 x10^-10^ | 17.49 | 8.06 | 34.94 |
| NMD-escape PTV | *ABCC8* | 0 | 6 | 1 | 0 | 0 | 51.28 |
|  | *HNF1B* | 0 | 1 | 1 | 0 | 0 | 2273.75 |
|  | *INS* | 3 | 1 | 1.70 x10^-5^ | 180.88 | 14.57 | 8809.85 |
|  | *KCNJ11* | 0 | 3 | 1 | 0 | 0 | 146.11 |
|  | *NEUROD1* | 8 | 23 | 2.80x10^-8^ | 21.02 | 8.13 | 48.72 |
|  | *PDX1* | 5 | 17 | 1.97 x 10^-5^ | 18.53 | 5.34 | 52.33 |
|  | *RFX6* | 1 | 5 | 0.09 | 12.09 | 0.26 | 108.27 |
| NMD-triggering synonymous | *ABCC8* | 19 | 923 | 0.36 | 1.24 | 0.75 | 1.96 |
|  | *HNF1B* | 3 | 281 | 0.64 | 0.64 | 0.13 | 1.9 |
|  | *INS* | 0 | 47 | 1 | 0 | 0 | 4.94 |
|  | *PDX1* | 2 | 93 | 0.67 | 1.3 | 0.16 | 4.83 |
|  | *RFX6* | 4 | 246 | 1 | 0.98 | 0.27 | 2.55 |
| NMD-escape synonymous | *ABCC8* | 0 | 19 | 1 | 0 | 0 | 12.95 |
|  | *HNF1B* | 0 | 0 | 1 | 0 | 0 | Inf |
|  | *INS* | 0 | 23 | 1 | 0 | 0 | 10.52 |
|  | *KCNJ11* | 1 | 197 | 0.39 | 0.31 | 0.008 | 1.73 |
|  | *NEUROD1* | 2 | 122 | 1 | 0.99 | 0.12 | 3.66 |
|  | *PDX1* | 1 | 85 | 1 | 0.71 | 0.02 | 4.07 |
|  | *RFX6* | 1 | 22 | 0.31 | 2.75 | 0.07 | 16.99 |

**Supplementary table 6: Sensitivity analysis gene burden test (MAF < 0.00005) in MODY cohort (n=2,571) with UK biobank (n=155,501) as controls**

**Supplementary table 7: Sensitivity analysis gene burden test (MAF < 0.0002) in MODY cohort (n=2,571) with UK biobank (n=155,501) as controls**

| Variant type | Gene | Allele count in MODY cohort | Allele count in population cohort  (UK biobank) | P value | Odds ratio | 95% CI lower limit | 95% CI upper limit |
| --- | --- | --- | --- | --- | --- | --- | --- |
| **NMD-triggering** **PTV** | *ABCC8* | 6 | 131 | 0.03 | 2.77 | 0.99 | 6.19 |
|  | *HNF1B* | 3 | 0 | 4.31x10^-6^ | Inf | 24.98 | Inf |
|  | *INS* | 0 | 2 | 1 | 0 | 0 | 322.22 |
|  | *PDX1* | 1 | 1 | 0.03 | 62.94 | 0.8 | 4940.62 |
|  | *RFX6* | 11 | 38 | 3.55 x10^-10^ | 17.49 | 8.06 | 34.94 |
| **NMD-escape PTV** | *ABCC8* | 0 | 6 | 1 | 0 | 0 | 51.28 |
|  | *HNF1B* | 0 | 1 | 1 | 0 | 0 | 2273.75 |
|  | *INS* | 3 | 1 | 1.70x x10^-5^ | 180.88 | 14.57 | 8809.85 |
|  | *KCNJ11* | 0 | 3 | 1 | 0 | 0 | 146.11 |
|  | *NEUROD1* | 8 | 23 | 2.80x10^-8^ | 21.02 | 8.13 | 48.72 |
|  | *PDX1* | 6 | 37 | 0.0001 | 10.22 | 3.52 | 24.42 |
|  | *RFX6* | 1 | 5 | 0.09 | 12.09 | 0.26 | 108.27 |
| **NMD-triggering** **synonymous** | *ABCC8* | 21 | 1295 | 1 | 0.98 | 0.6 | 1.51 |
|  | *HNF1B* | 5 | 481 | 0.37 | 0.63 | 0.2 | 1.48 |
|  | *INS* | 0 | 47 | 1 | 0 | 0 | 4.94 |
|  | *PDX1* | 2 | 93 | 0.67 | 1.3 | 0.16 | 4.83 |
|  | *RFX6* | 10 | 656 | 1 | 0.92 | 0.44 | 1.71 |
| **NMD-escape synonymous** | *ABCC8* | 0 | 80 | 0.64 | 0 | 0 | 2.85 |
|  | *HNF1B* | 0 | 0 | 1 | 0 | 0 | Inf |
|  | *INS* | 0 | 69 | 0.63 | 0 | 0 | 3.32 |
|  | *PDX1* | 1 | 147 | 0.74 | 0.41 | 0.01 | 2.33 |
|  | *KCNJ11* | 5 | 368 | 0.84 | 0.82 | 0.26 | 1.94 |
|  | *NEUROD1* | 4 | 218 | 0.79 | 1.11 | 0.30 | 2.88 |
|  | *RFX6* | 1 | 22 | 0.31 | 2.75 | 0.07 | 16.99 |

**Supplementary table 8: Sensitivity analysis gene burden test for all PTVs (no MAF threshold) in MODY cohort (n=2,571) with UK biobank (n=155,501) as controls**

| Variant type | Gene | Allele count in MODY cohort | Allele count in population cohort  (UK biobank) | P value | Odds ratio | 95% CI lower limit | 95% CI upper limit |
| --- | --- | --- | --- | --- | --- | --- | --- |
| **NMD-triggering** **PTV** | *ABCC8* | 6 | 131 | 0.03 | 2.77 | 0.99 | 6.19 |
|  | *HNF1B* | 3 | 0 | 4.31E-06 | Inf | 24.98 | Inf |
|  | *INS* | 0 | 2 | 1 | 0.00 | 0.00 | 322.22 |
|  | *PDX1* | 1 | 1 | 0.03 | 62.94 | 0.8 | 4940.62 |
|  | *RFX6* | 11 | 38 | 3.55E-10 | 17.49 | 8.06 | 34.94 |
| **NMD-escape PTV** | *ABCC8* | 0 | 6 | 1 | 0.00 | 0.00 | 51.28 |
|  | *HNF1B* | 0 | 1 | 1 | 0.00 | 0.00 | 2273.75 |
|  | *INS* | 3 | 1 | 1.70E-05 | 180.88 | 14.57 | 8809.85 |
|  | *KCNJ11* | 0 | 3 | 1 | 0 | 0 | 146.11 |
|  | *NEUROD1* | 8 | 23 | 2.80x10^-8^ | 21.02 | 8.13 | 48.72 |
|  | *PDX1* | 6 | 37 | 0.0001 | 10.22 | 3.52 | 24.42 |
|  | *RFX6* | 1 | 5 | 0.09 | 12.09 | 0.26 | 108.27 |

**Supplementary table 9: Sensitivity analysis gene burden test (MAF < 0.0001) in MODY cohort (n=2,571) gnomAD v3.1.1 (n=34,029) as controls**

| Variant type | Gene | Allele count in MODY cohort | Allele count in population cohort  (gnomAD) | P value | Odds ratio | 95% CI lower limit | 95% CI upper limit |
| --- | --- | --- | --- | --- | --- | --- | --- |
| **NMD-triggering PTV** | *ABCC8* | 6 | 26 | 0.02 | 3.05 | 1.03 | 7.59 |
|  | *GCK* | 126 | 5 | 3.62x10^-109^ | 72.06 | 43.23 | 127.67 |
|  | *HNF1A* | 74 | 3 | 2.37 x10^-70^ | 225.87 | 74.32 | 1120.23 |
|  | *HNF1B* | 14 | 2 | 7.63 x10^-15^ | 92.62 | 21.26 | 822.94 |
|  | *HNF4A* | 23 | 2 | 3.21 x10^-23^ | 127.82 | 31.57 | 1118.39 |
|  | *INS* | 0 | 1 | 1 | 0 | 0 | 511.79 |
|  | *PDX1* | 1 | 0 | 0.07 | Inf | 0 | Inf |
|  | *RFX6* | 9 | 9 | 1.14 x 10^-6^ | 13.23 | 4.65 | 37.60 |
| **NMD-escape PTV** | *ABCC8* | 0 | 0 | . | . | . | . |
|  | *GCK* | 11 | 0 | 4.77 x10^-12^ | Inf | 27.79 | Inf |
|  | *HNF1A* | 11 | 1 | 1.03 x10^-10^ | 99.62 | 14.47 | 4287.16 |
|  | *HNF1B* | 0 | 0 | . | . | . | . |
|  | *HNF4A* | 2 | 2 | 0.04 | 11.08 | 0.80 | 152.82 |
|  | *INS* | 3 | 0 | 0.0003 | Inf | 5.47 | Inf |
|  | *KCNJ11* | 0 | 2 | 1 | 0 | 0 | 70.39 |
|  | *NEUROD1* | 8 | 6 | 1.22 x10^-6^ | 17.63967 | 5.36571 | 61.70326 |
|  | *PDX1* | 5 | 5 | 2.21 x 10^-7^ | 62.99 | 14.49 | 273.78 |
|  | *RFX6* | 1 | 6 | 0.4 | 2.20 | 0.05 | 18.18 |
| **NMD-triggering synonymous** | *ABCC8* | 17 | 260 | 0.6 | 0.86 | 0.49 | 1.41 |
|  | *GCK* | 7 | 70 | 1.00 | 0.97 | 0.38 | 2.1 |
|  | *HNF1A* | 17 | 120 | 0.3 | 1.28 | 0.72 | 2.14 |
|  | *HNF1B* | 4 | 68 | 0.82 | 0.78 | 0.21 | 2.08 |
|  | *HNF4A* | 9 | 79 | 0.4 | 1.26 | 0.56 | 2.52 |
|  | *INS* | 0 | 8 | 1.00 | 0.00 | 0.00 | 7.75 |
|  | *PDX1* | 2 | 24 | 0.70 | 1.10 | 0.13 | 4.44 |
|  | *RFX6* | 8 | 98 | 0.9 | 1.08 | 0.45 | 2.21 |
| **NMD-escape synonymous** | *ABCC8* | 0 | 10 | 1.00 | 0.00 | 0.00 | 5.90 |
|  | *GCK* | 3 | 12 | 0.2 | 2.42 | 0.44 | 8.97 |
|  | *HNF1A* | 3 | 12 | 0.2 | 2.26 | 0.41 | 8.38 |
|  | *HNF1B* | 0 | 1 | 1.00 | 0.00 | 0.00 | 511.84 |
|  | *HNF4A* | 1 | 13 | 1.00 | 0.85 | 0.02 | 5.67 |
|  | *INS* | 0 | 12 | 1.00 | 0.00 | 0.00 | 4.76 |
|  | *KCNJ11* | 2 | 70 | 0.24 | 0.38 | 0.04 | 1.42 |
|  | *NEUROD1* | 3 | 59 | 0.80 | 0.67 | 0.13 | 2.06 |
|  | *PDX1* | 1 | 25 | 1.00 | 0.53 | 0.01 | 3.23 |
|  | *RFX6* | 1 | 4 | 0.3 | 3.31 | 0.07 | 33.41 |

2,571 individuals had tNGS. 2,600 individuals underwent Sanger sequencing for either ***GCK*** (n = 941), ***HNF1A*** (n = 1,292), or ***HNF4A*** (n = 602).

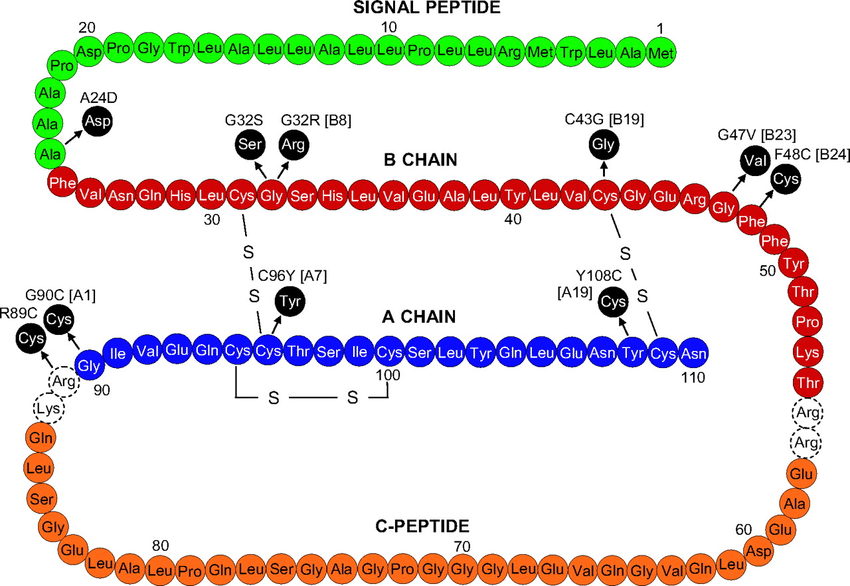

**p.Gln78***

**p.Leu82GlyfsTer52**

**p.Glu83ValfsTer58**

**p.Cys95***

S

S

**p.Glu57***

**p.Gly69Alafs*62**

**p.Gly71Alafs*60**

**p.Gln78Argfs*53**

**p.Cys96Valfs*35**

**B CHAIN**

**p.Gly73Trpfs?**

**Supplementary figure 1: Figure showing NMD-escape PTVs in *INS.*** The structure of preproinsulin is shown in the figure, with signal peptide in green, B chain in red, C peptide in orange and A chain in blue, with 3 disulphide bonds shown. Highlighted variants are seen in the MODY cohort, others reported in literature. (Figure adapted from https://doi.org/10.1016/j.molmet.2021.101280)

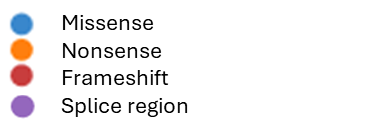

a) ***GCK***

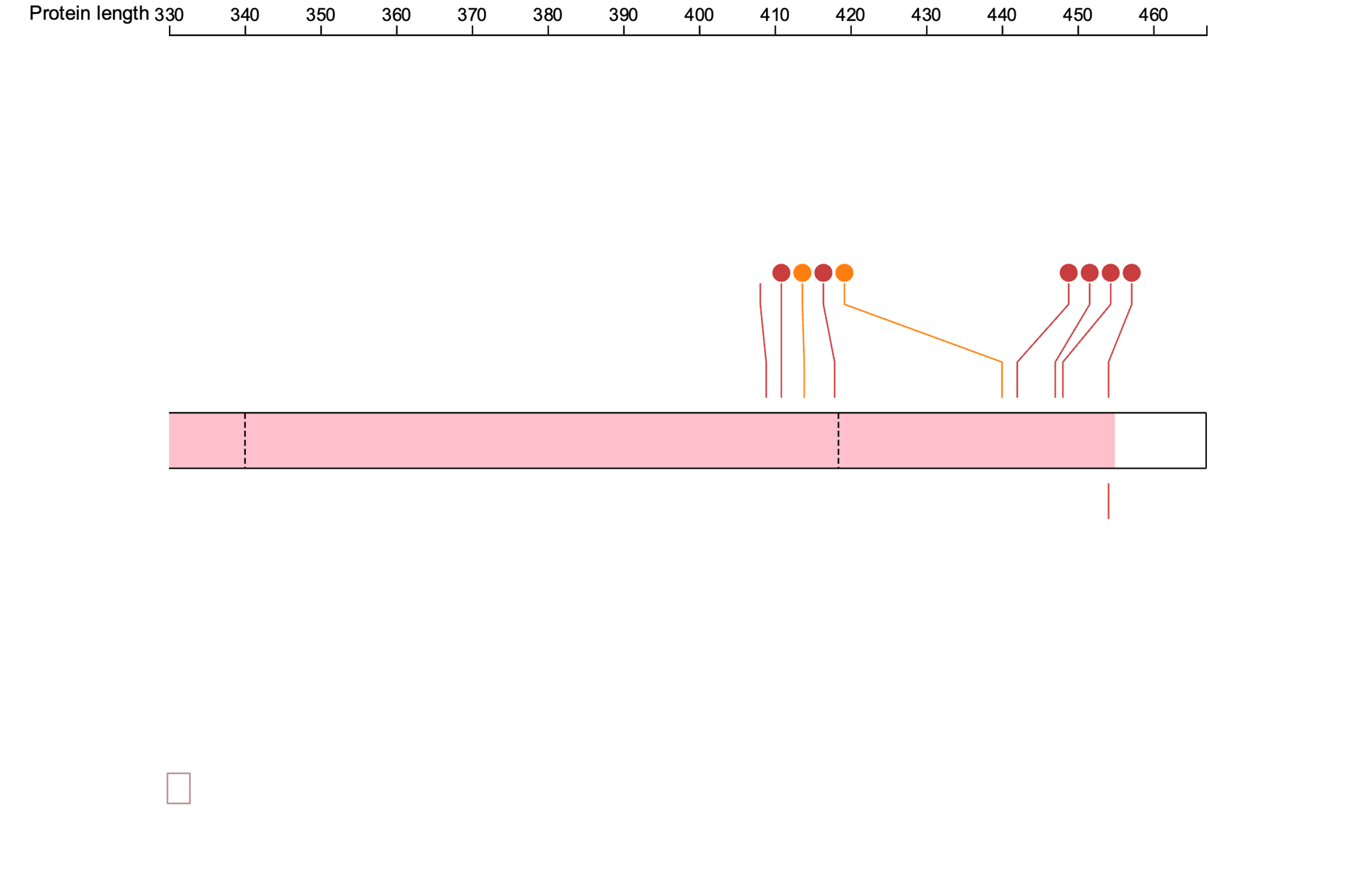

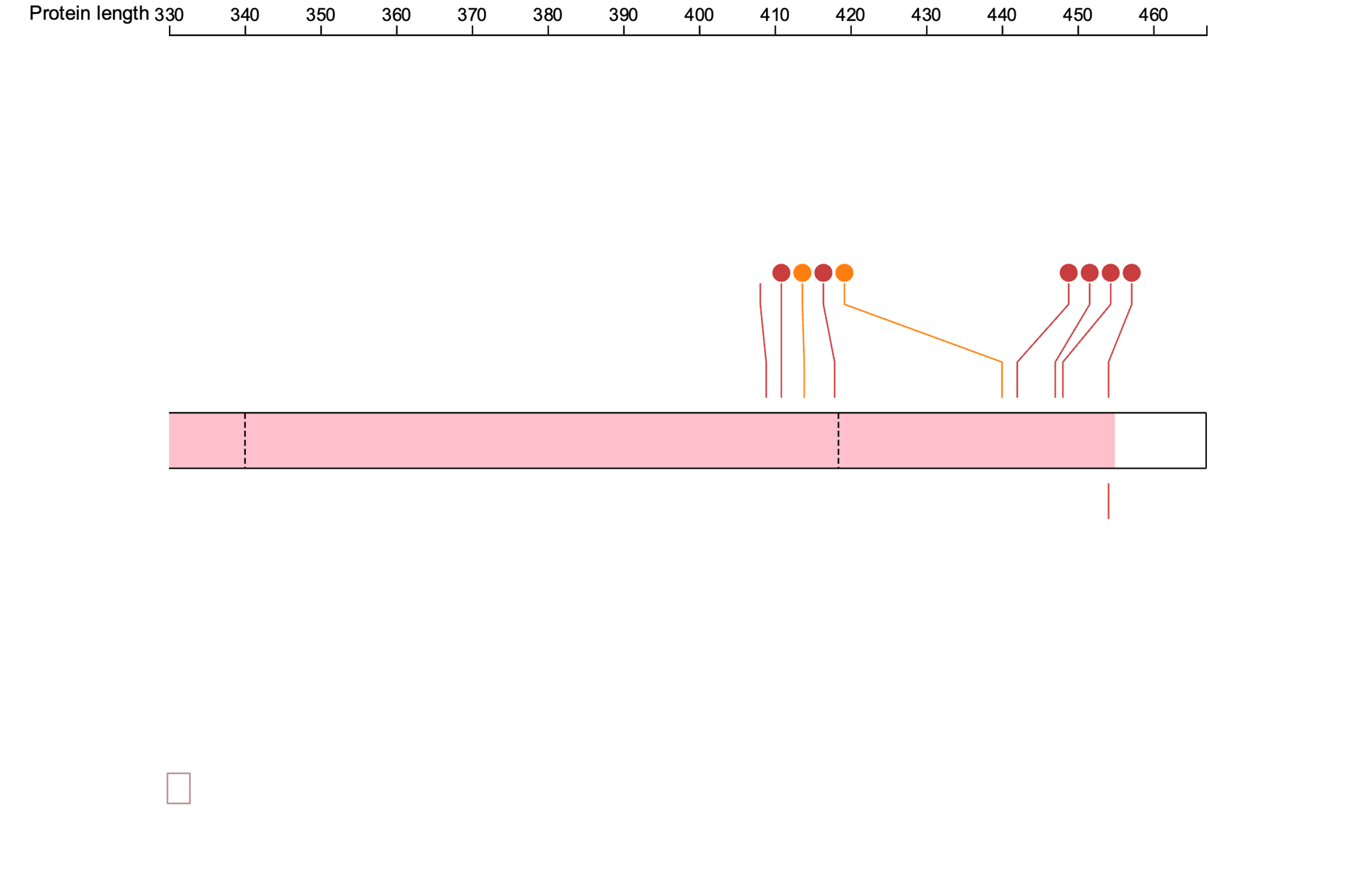

b) ***HNF1A***

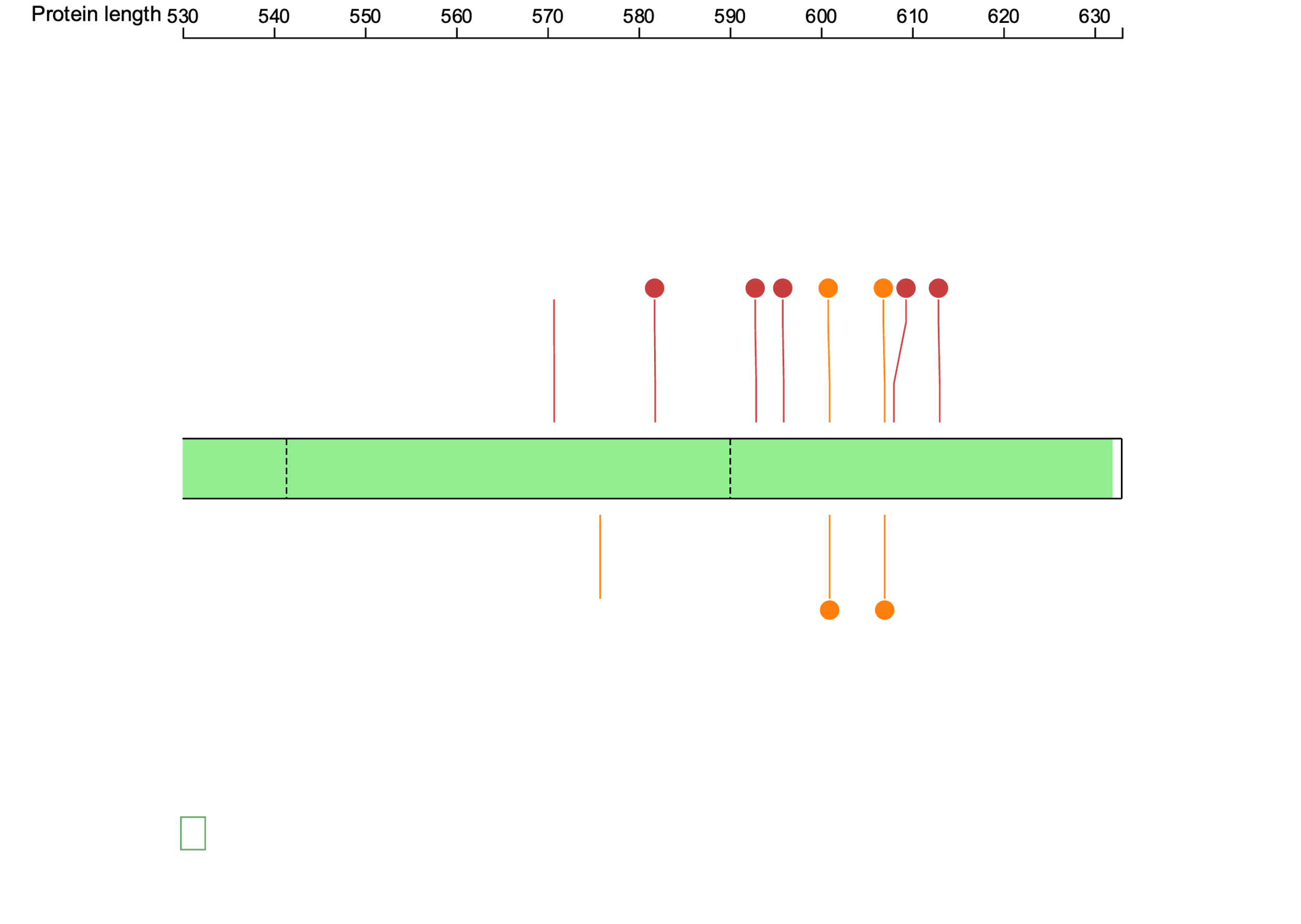

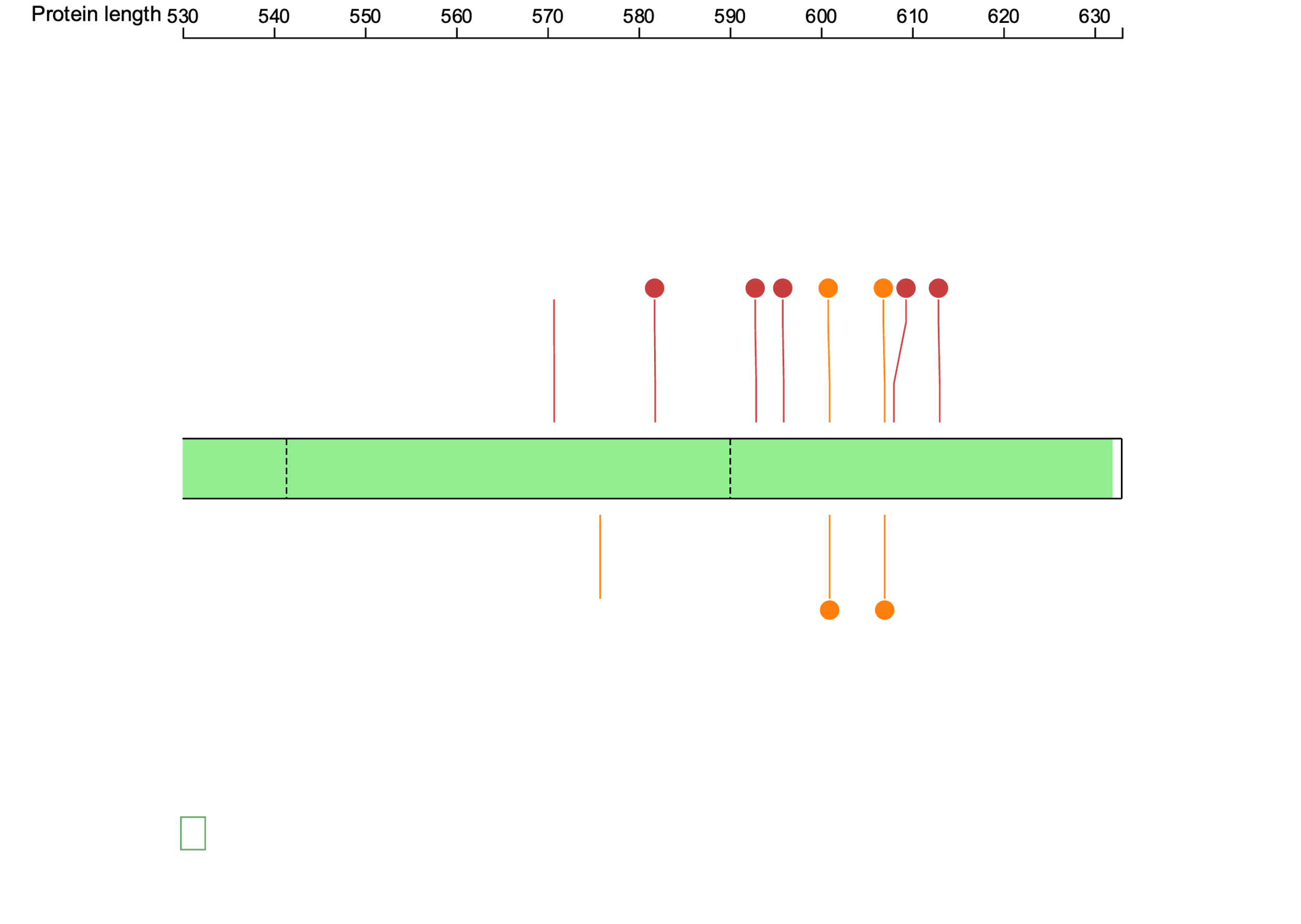

c) ***HNF4A***

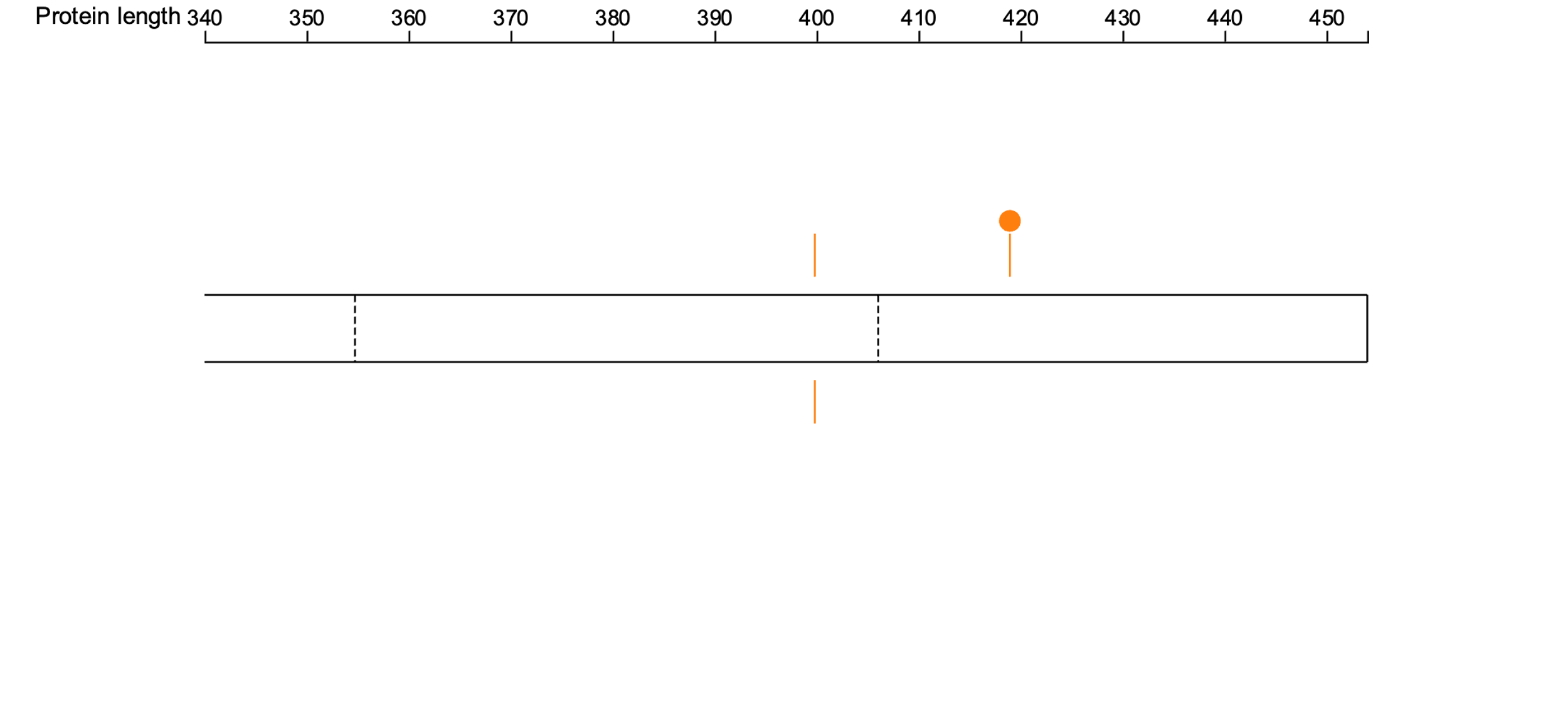

d)
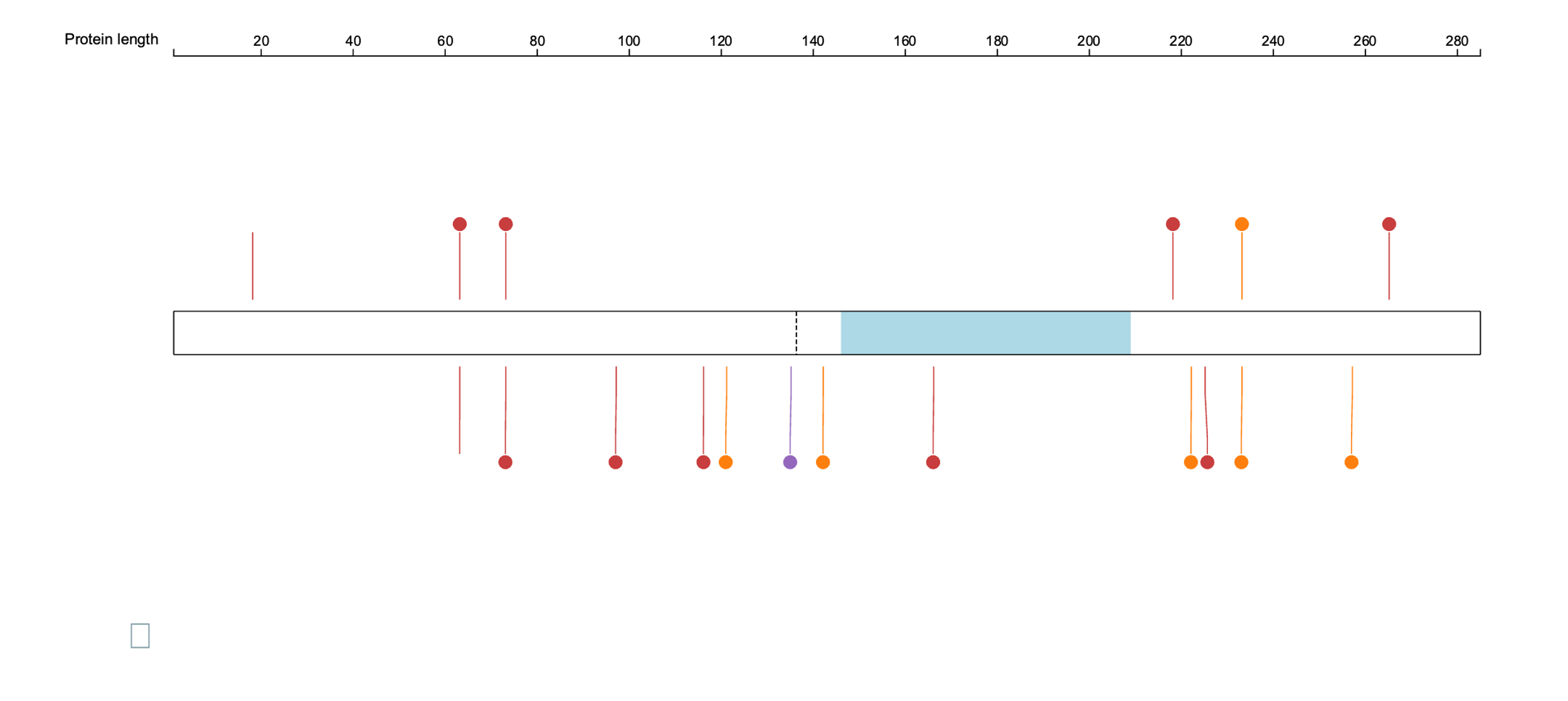
***PDX1***

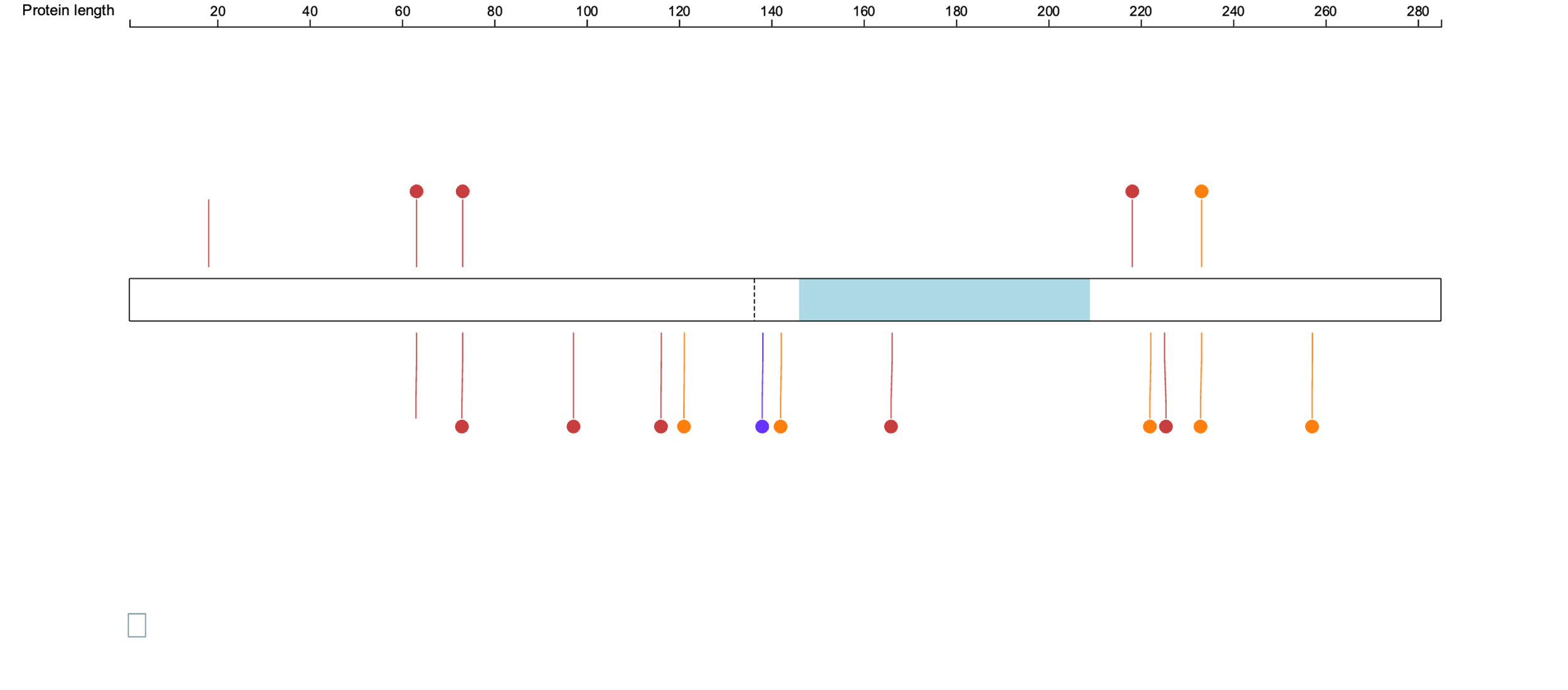

e) ***RFX6***

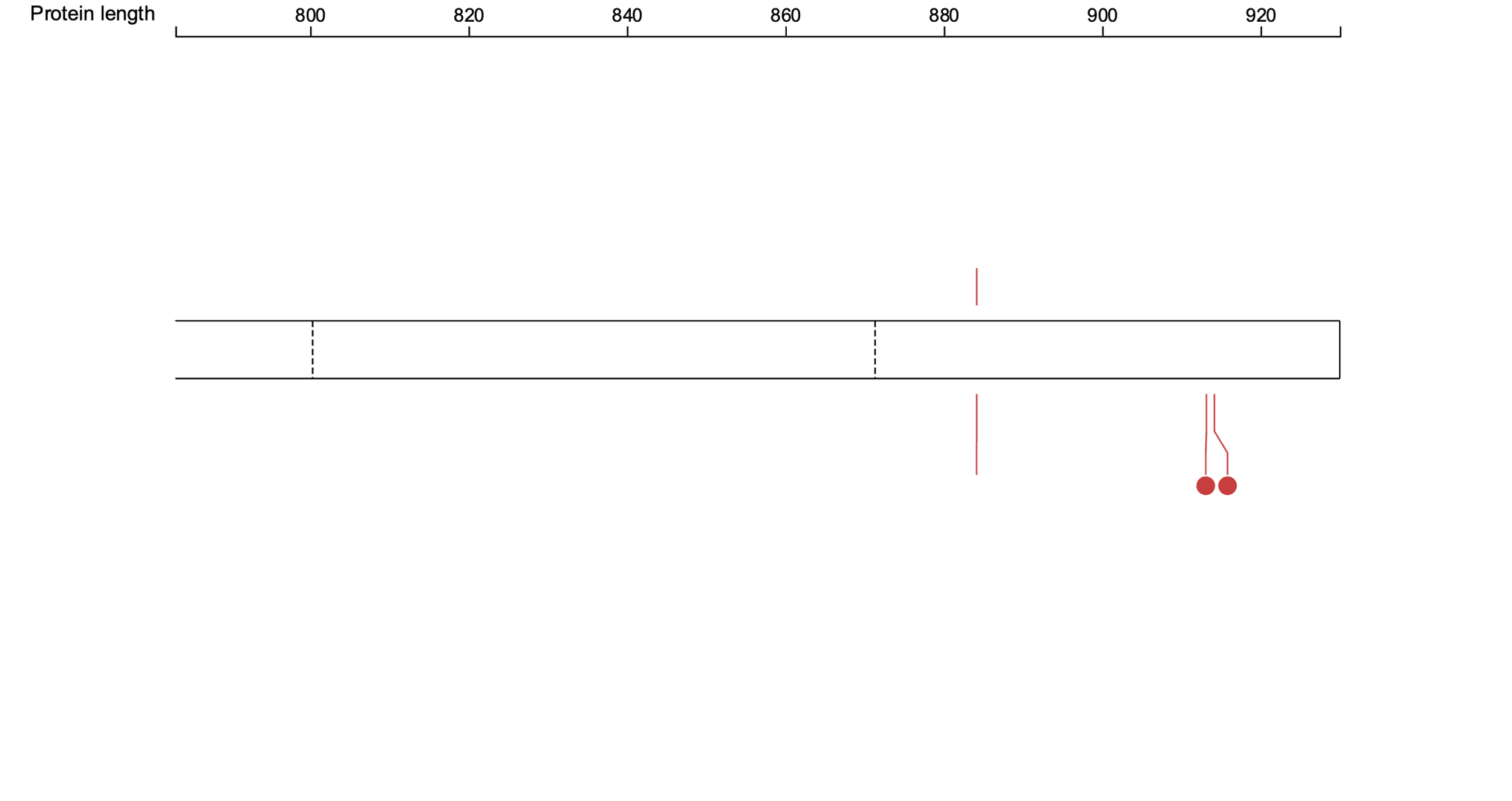

**Supplementary figure 2: Figure showing NMD-escape PTVs in MODY genes.** This includes NMD-escape LOF variants split into nonsense, frameshift and splice region variants. The functional domains within each gene have been indicated. Variants identified in the MODY cohort are shown above the gene, while those from the UK Biobank are displayed below. Figures were generated using ProteinPaint (<https://proteinpaint.stjude.org/>).

**Supplementary table 10: Case reports published about *INS* NMD-escape LoF variants in young-onset diabetes**

| Variant | Ref | Study type | No. of families | No. of cases with variant/No. of cases with diabetes | Age of diagnosis | Sex | Treatment | LOD score | Notes |
| --- | --- | --- | --- | --- | --- | --- | --- | --- | --- |
| p.Glu57* | PMID: 34362814 | Cohort | 1 | 1/1 | Not available | Not available | Not available | n/a | - |
| p.Gly69Aalfs*62 | PMID: 38553172 | Case report | 1 | 4/4 | 10-25 | 3 F, 1 M | Metformin + Gliclazide + FGM, CSII MM780G, Insulin + FGM, Sitagliptin/Metformin + Insulin. | n/a | All individuals negative for all antibodies tested. |
| p.Gly71Alafs*60 | PMID: 36504295 | Cohort | 1 | 1/1 | 5-10 | M | Not available | n/a | Proband has no affected parents. |
| p.Gly73Trpfs*? | PMID: 30182532 | Case report | 1 | 5/7 | 15-40, ages not available for five individuals | 4 F, 3 M | Insulin + Metformin, Insulin, Gliclazide + Metformin, Dialysis + No Diabetes meds, Gliclazide + Metformin. | 1.2 | No diabetes in 2 carriers- one aged 80-90, one aged 5-10. |
| p.Gln78Argfs*53 | PMID: 25721872 | Case report | 1 | 3/3 | 10-30 | 1 F, 2 M | Insulin, Diet, Metformin + Insulin + Detemir + Sitagliptin + Glimepirid. | n/a | Proband- GAD, IA2 negative. |
| p.Cys96Valfs*35 | PMID: 32086287 | Cohort | 1 | 1/1 | Not available | Not available | Not available | n/a | - |

**Supplementary table 11: List of *INS* missense variants in patients used for comparison with patients with NMD-escape PTVs in *INS***

| Variant | Number of individuals |
| --- | --- |
| p.R6C | 1 |
| p.A24D | 1 |
| p.A24V | 1 |
| p.H29Q | 2 |
| p.L30V | 1 |
| p.G32R | 1 |
| p.G32S | 3 |
| p.C43F | 2 |
| p.G44R | 1 |
| p.R46Q | 9 |
| p.R55C | 9 |
| p.S85C | 2 |
| p.R89C | 5 |
| p.C96S | 1 |
| p.C96Y | 4 |

**Supplementary table 12: Gene and protein predictive scores for our genes of interest**

| **Gene** | **pLI** | **LOUEF** | **sHet** | **pHaplo** | **pTriplo** | **Missense Z score** | **pDN** | **pGOF** | **pLOF** |
| --- | --- | --- | --- | --- | --- | --- | --- | --- | --- |
| *GCK* | 0 | 0.87 | 0.013 | 0.71 | 0.98 | 2.79 | 0.556 | 0.581 | 0.36 |
| *HNF1A* | 0.99 | 0.48 | 0.056 | 0.92 | 0.78 | 2.09 | 0.385 | 0.311 | 0.727 |
| *HNF4A* | 1 | 0.45 | 0.043 | 0.55 | 0.88 | 2.71 | 0.61 | 0.476 | 0.51 |
| *HNF1B* | 1 | 0.24 | 0.2775 | 0.96 | 0.96 | 2.08 | 0.353 | 0.267 | 0.815 |
| *INS* | 0.02 | 1.2 | 0.082 | 0.79 | 0.73 | 0.86 | 0.855 | 0.845 | 0.351 |
| *RFX6* | 0 | 0.69 | 0.011 | 0.55 | 0.48 | 0.92 | 0.64 | 0.423 | 0.436 |
| *PDX1* | 0 | 1.64 |  | 0.71 | 0.52 | -1.15 | 0.668 | 0.598 | 0.49 |
| *NEUROD1* | 0.42 | 0.69 | 0.037 | 0.92 | 0.95 | 1.05 | 0.513 | 0.222 | 0.728 |
| *ABCC8* | 0 | 0.83 | 0.008 | 0.59 | 0.47 | 2.85 | 0.777 | 0.803 | 0.219 |
| *KCNJ11* | 0 | 1.07 | 0.026 | 0.5 | 0.78 | 2.86 | 0.806 | 0.874 | 0.219 |

Gene predictive scores:

pLI: Probability of Loss-of-function Intolerance. (closer to 1, more intolerant of LoF mutations). LOEUF: Loss-of-function Observed / Expected Upper bound Fraction. (closer to 0, more intolerant of LoF mutations). sHet: Selection coefficient of heterozygous loss-of-function variants (Higher numbers, more dosage intolerant). pHaplo: Predicted Probability of Haploinsufficiency (higher numbers, more dosage sensitive). pTriplo: Predicted Probability of Triplosensitivity (higher numbers, more dosage sensitive). Missense Z score: Missense Z scores represent the deviation of observed missense variant counts from the expected number in gnomAD (>3.09 - significantly constrained. Positive scores- more likely to be intolerant to missense variation).

Protein predictive scores:

pDN: Dominant-negative mechanism propensity of the protein. (Higher scores- genes likely to be associated with DN disease mechanism). pGOF: Gain-of-function mechanism propensity of the protein (Higher scores- genes likely to be associated with GoF disease mechanism). pLOF: Loss-of-function mechanism propensity of the protein (Higher scores- genes likely to be associated with LoF disease mechanism)

**Supplementary table 13: Power calculations**

| Genes | Cohort | MAF | Power | Minimum detectable odds ratio |
| --- | --- | --- | --- | --- |
| *ABCC8*, *HNF1B*, *INS*, *NEUROD1*, *KCNJ11*, *PDX1*, *RFX6* | tNGS (n=2,571) | 0.0001 | 0.8 | 9.8 |
| *GCK* | Sanger+tNGS (n=3,512) | 0.0001 | 0.8 | 8.3 |
| *HNF1A* | Sanger+tNGS (n=3,863) | 0.0001 | 0.8 | 7.9 |
| *HNF4A* | Sanger+tNGS (n=3,073) | 0.0001 | 0.8 | 8.9 |

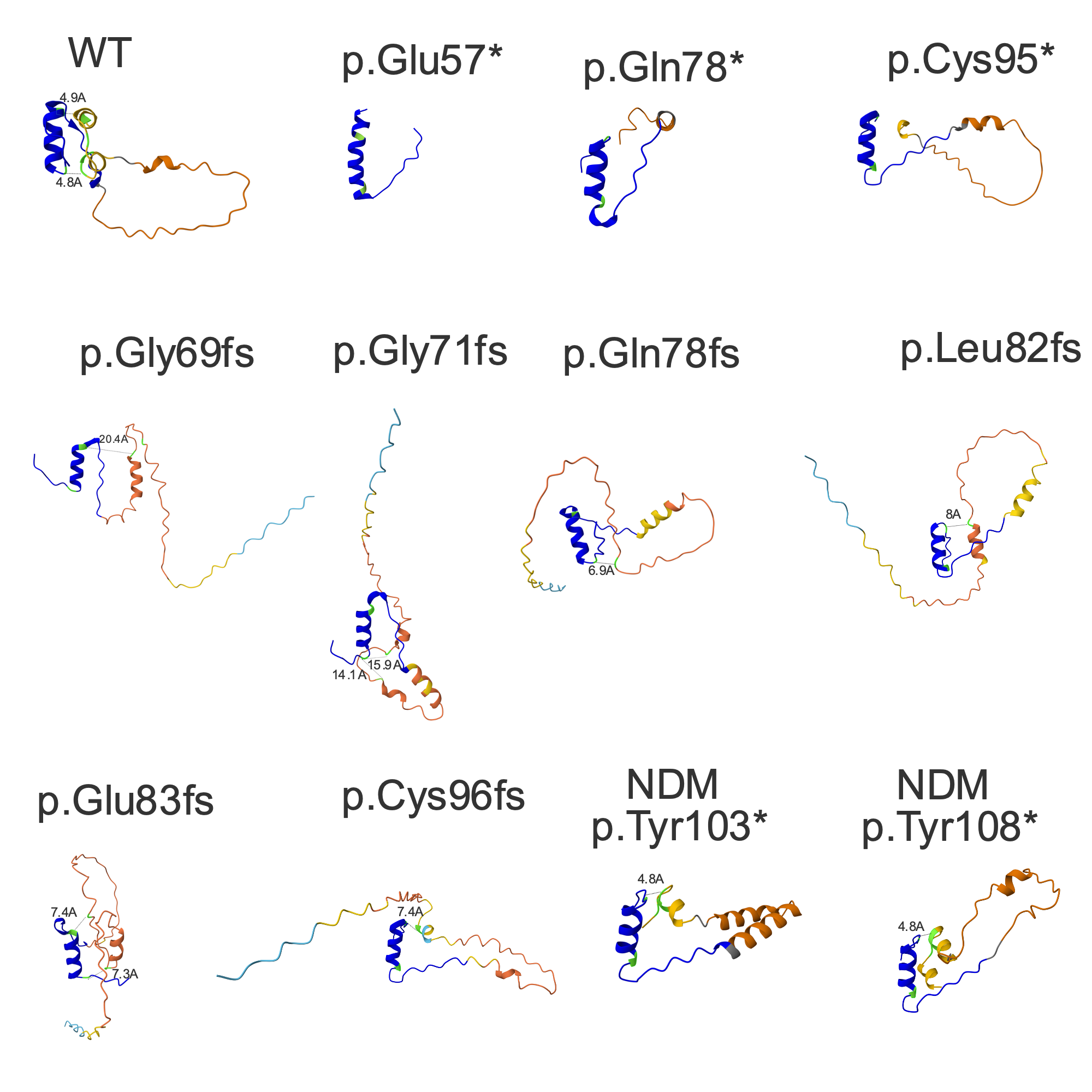

**Supplementary figure 3: Protein predictions for *INS* NMD-escape.** Proteins were modelled using Alphafold. B chains are shown in dark blue. Cysteines are shown in green and predicted distances between cysteines are measured in Angstrom and shown in the plots.
